# Identifying barriers and potential solutions to improve equitable access to community eye services in central Kenya: a rapid exploratory sequential mixed methods study

**DOI:** 10.1101/2024.03.13.24304156

**Authors:** Luke Allen, Sarah Karanja, Michael Gichangi, Cosmas Bunywera, Emmaculate Muturi, Dickson Gachobi, Purity Kathure, Elizabeth Mutile Muasa, Lorna Mutwiri, Lorna Kajuju, Faith Kagwiria, Benjamin Ntabathia, Hillary Rono, David Macleod, Min Jung Kim, Malebogo Tlhajoane, Matthew J. Burton, Jacqueline Ramke, Nigel M. Bolster, Andrew Bastawrous

## Abstract

**Background:** Recent research has found that less than half of people identified with an eye problem in Meru county’s screening programme were able to access care, with younger adults being the least likely to receive the care they needed. We aimed to interview and survey members of this ‘left-behind’ group to explore barriers and identify potential solutions using a rapid mixed-methods approach.

**Methods:** First, we conducted interviews to explore perceptions of barriers and potential solutions. Next, we asked a representative sample to rank the suggested solutions by likely impact. Finally, we held a multistakeholder meeting to identify which of the top-ranked interventions offered the best balance of impact, feasibility, cost, and potential risks. We used a deductive matrix and thematic analysis to rapidly analyse the interview data.

**Results:** We conducted 67 interviews. Barriers to access included long queues, conflicting work engagements, and lack of clear information. Proposed solutions focused on reducing queue lengths, providing better counselling and clinic information, holding mop-up clinics, and maintaining adequate stocks & supplies. We conducted ranking surveys with 401 additional people from the left-behind group. All proposed solutions were ranked at moderately-to-highly likely to improve equitable access. Fifteen people attended the multistakeholder meeting, including community representatives. Workshop participants unanimously selected enhanced counselling and SMS reminders as the interventions that offered the best balance of impact, risk, cost, and feasibility. The other proposed solutions were deemed impractical or unaffordable.

**Conclusion:** Rapid mixed-methods and multistakeholder collaboration were used to identify a range of potential service modifications that will be implemented within the ongoing programme. Our approach was centred on the experiences and perceptions of those who face the highest barriers to care.

**Research in Context:** *Evidence before this study:* Previous research in Kenyan community screening programmes has shown that at least half of those found to have an eye health need will not be able to access care at their local treatment clinic, even if the care is provided free. Work in Meru County has shown that younger adults less are likely than any other sociodemographic group to check-in at their local clinic, but it’s not clear what the specific barriers are for this group. Across the African continent, approximately half of all ambulatory appointments are missed across all specialities, and sociodemographic inequalities are ubiquitous. In pursuit of Universal Health Coverage (UHC) and the Primary Health Care principles of equity and justice, health system managers are increasingly focused on identifying, trying to understand, and then address unequal access to care, however the traditional approach to identifying barriers and solutions has tended to centre around expert opinion rather than engagement with affected groups.

*Added value of this study:* This study builds on previous efforts to introduce routine sociodemographic data collection into the county-wide eye screening programme operating in Meru, Kenya, as well as additional sites in Meru County, Botswana, Nepal, and Uttar Pradesh. Having already identified younger adults as the least likely to receive care in Meru County, this study introduces a novel mixed-methods approach for engaging with members of this left-behind group to rapidly identify barriers and scalable solutions. We used innovative methods to complete interviews and qualitative analysis in under two weeks, followed by a rapid survey to rank the potential solutions that emerged from this work with a representative sample of younger adults who had not been able to access care. Finally, a multistakeholder workshop with strong local and lay representation identified the top-ranked solutions that would be feasible to introduce and test within the ongoing screening programme. In addition to local evidence for action, this study presents an approach that any community-based programme could use to generate robust, non-tokenistic insights from affected communities within a matter of weeks, minimising the research time requirement and number of senior researchers required whilst maintaining rigorous scientific standards.

*Implications of all the available evidence:* Equitably advancing UHC is predicated on identifying and overcoming unique barriers to care, however existing efforts rarely involve consultation or co-creation with affected communities. Building on existing rapid qualitative and mixed-methods methods, we have developed a cutting-edge approach to identify barriers, prioritise solutions, and identify service modifications that are feasible to introduce. We have applied this approach in Meru County, where younger adults – who were the least likely to access care – suggested a bundle of interventions centring on improving the provision of information and SMS reminders. Our research group will use an embedded RCT to implement and test this bundle, in the context of an equity-focused continuous improvement model that we are also implementing in Botswana, India and Nepal to incrementally improve access for all, with a focus on left-behind groups.

## Introduction

Improving equitable access to community health services lies at the heart of Universal Health Coverage (UHC) and ‘leaving no one behind’ is the ‘central, transformative promise’ of the Sustainable Development Goals.^1–4^ WHO’s *Thirteenth General Programme of Work* states that ‘the main challenge to making progress towards UHC comes from persistent barriers to accessing health services’.

Our research collaborative is developing and testing a novel approach to identify and address inequitable access to care using the ‘IM-SEEN’ approach (‘<u>Im</u>provement <u>s</u>tudies for <u>e</u>vidence-based and <u>e</u>quitable innovatio<u>n</u>’). This involves identifying which groups are being left behind in a given programme; engaging with these groups to understand the unique barriers they face and their ideas for service improvements; and then testing these potential solutions with embedded randomised controlled trials (RCTs).^5^

We are applying this model in the context of community-based eye screening programmes in Botswana, India, Kenya, and Nepal. Uncorrected visual impairment affects over a billion people worldwide, levying major social and economic costs, despite the availability of highly cost-effective treatments like spectacles and cataract surgery.^6^ Our first set of findings from a cross-sectional equity analysis of over 4,000 people in Kenya’s Meru county found that only 46% of those found to have an eye need were able to access their free local treatment outreach clinic.^7^ We found that younger adults, males, and those working in sales, services, or manual jobs were the least likely to receive the care they need. Age was the strongest predictor of poor access, with less than a third of people aged 18-44 years receiving care compared to two thirds of those aged >45 years, even after controlling for severity of eye condition and a wide range of other factors.

Traditionally, ideas for how to improve programmes come from ‘experts’, service providers, or surveys of service users - rather than affected people themselves.^8,9^ In the context of renewed interest in Primary Health Care^10–12^ and the insidious persistence of colonialism and epistemic injustice in global health,^13–15^ increasing attention is being paid to person- and community-centredness. Simply put, advancing equitable access to health services must be done *with*, rather than *to,* or *on behalf of* left-behind groups.^9^

In this study we aimed to engage with younger adults who had not been able to access eye care in Meru in order to explore their perceptions of how the local services could be modified to improve access. Working within a live programme, we aimed to deliver robust, non-tokenistic, and generalisable findings within a matter of weeks, with a view to testing suggested service modifications with a subsequent embedded RCT.

## Methods

### Setting

Meru is a county with a population of 1.5 million in central Kenya, 110 miles north of Nairobi. It includes Mount Kenya and Meru National Park. The capital, Meru town, is home to a quarter of a million people. Agriculture is the main source of employment, with khat and tea representing important cash crops. Kenya’s Vision Impact Programme (‘VIP’) has been operating in Meru since July 2022, and has reached over 350,000 people to date, according to internal data. Teams of screeners go house-to-house testing all adults’ vision using a simple smartphone-based app developed by Peek Vision.^16^ Screeners refer people whose visual acuity falls below 6/12; those who have a red eye or another issue upon basic visual inspection; and anyone who feels they have an eye problem, even if there are no clinical signs and their visual acuity is >6/12. Our research team has been working with screeners to gather sociodemographic data from every person who screened positive and was referred to an outreach clinic for further assessment and treatment between April – July 2023. As stated above, we had previously found that younger adults are the least likely to be checked-in at treatment clinics but we did not know what the main barriers were or what could be done about them.

### Research paradigm, theory, and methodology

We used a pragmatist philosophical paradigm^17,18^ and a phenomenological approach^19,20^ to explore these young adults’ lived experiences and perceptions of barriers to accessing eye clinics, and potential solutions. We grounded our work in the complementary frameworks developed by Levesque et al and Obrist et al.^21,22^ Both conceptualise access to care in terms of service and service-user characteristics. This distinction is helpful as our ultimate aim was to identify service modifications that improve accessibility for younger adults. We required mixed methods to answer a multi-layered question: what are the main barriers to accessing eye services in each location and what could be done about them?

### Methods overview

This study was conducted in three stages. In Stage 1, we used interviews to generate a long-list of perceived barriers and potential solutions. Then, in Stage 2, to move from subjective experiences to generalisable service modifications, we conducted a telephone survey where a representative sample of younger adults who did not receive care ranked each of the suggested solutions by likely impact. Finally, in Stage 3, these ranked solutions were reviewed by a multistakeholder group who identified a package of interventions to test based on likely impact, feasibility, cost, and risks.

### Team composition and reflexivity

This project was part of the broader ‘IM-SEEN’ programme of work that seeks to develop a new, rapid, robust, and responsive approach to continuously improving access to care, starting in the field of community-based eye screening programmes in Botswana, India, Kenya, Nepal. LSHTM-based researchers (LA, AB, MB, JR, DM & MK) working with Kenya’s Ministry of Health eye lead MG, AB and NB from Peek Vision - the screening programme software provider, and SK - the local research lead SK based at KEMRI, had already conducted a collaborative equity assessment of Meru’s VIP programme. LA – a mid-career British clinician, policy advisor, and mixed-methods public health researcher - led the development of the methodological approach to be used in all countries to engage with members of the left behind groups. LA worked closely with SK – a mid-career female Kenyan public health social scientist – to tailor the approach for Meru County, supported by the wider team. LA and SK recruited and trained six local, early-career data collectors (DG, EMM, EM, PK, BN and FG) to conduct the interviews and surveys. We were interested in understanding the barriers and solutions as perceived and described by affected people in their own words. SK and LA facilitated the multistakeholder workshop where findings were interpreted by lay representatives, other members of the left behind group, and local programme managers. This local multistakeholder group collectively made the final decisions on which suggested service modifications to take forward for implementation.

### Stage 1: interviews

#### Recruitment and sample size

Peek Vision – the programme software provider - provided us with a list of every person aged 18-44 years who had not been able to access their clinic appointment in Meru. In random order, we phoned people from this list to invite them to participate in the interviews, and sought recorded verbal informed consent. We tried each person three times before moving on to the next.

We planned to use Guest and colleagues’ approach to determine our sample size based on thematic saturation, using a ‘base’ of 12 interviews followed by runs of two interviews with a 0% new information threshold.^23^ In other words, we aimed to continue recruiting interviewees until no new themes emerged after two interviews in a row, with a minimum sample size of 14 (‘12+2’ approach).

#### Interview modality

We wanted to use telephone interviews, based on empirical evidence that they can be completed faster at lower cost than in-person interviews, and with equivalent data richness.^24–28^ However, we were not entirely convinced that the data would be equivalent. As such, we decided to recruit two separate samples and use both modalities, conducting an embedded mode-comparison study^29^ that will be reported elsewhere.

#### Data collection

Three pairs of Kenyan data collectors with at least basic qualitative training and fluency in English, Kiswahili, and the local dialect conducted semi-structured interviews using the topic guide summarised in Box 1 (see Appendix 1 for the full script). For the telephone interviews, calls were made on speakerphone in a private space and recorded using the phone’s inbuilt call recording app. As one data collector conducted the interview, the other noted down the times at which each unique barrier and potential solution was mentioned. After the call, the interview recording was immediately replayed and the data collectors entered verbatim quotes directly from the audio into our analytic matrix. The same process was used for in-person interviews, but with an audio recorder instead of a mobile phone. Our decision to use direct-from-audio transcription was based on findings from a background systematic review that we conducted on rapid qualitative approaches.^30^ Interviewees did not review their transcribed quotes in the matrix. In-person interviews were conducted in private rooms in four different health facilities where interviewees’ responses could not be overheard. Only the data collectors and the interviewee were present for each interview.

##### Box 1

###### Topic guide

Barriers

- In your own words, can you talk me through why we didn’t see you at that clinic?
Probing questions

Are there any other factors that prevented you from attending?
Is there anything else you’d like to share?
Solutions

- What would make the biggest difference in addressing these barriers? Probing questions
- else would help?
- other changes could we make to the programme that would make it easier for you to attend?
- there any other specific changes that we could make to the way that the programme or eye clinics run?
- mentioned [list their proposed solutions]. Some of these may be beyond our control, but if we managed to [list their proposed programme-related changes], do you think that would be enough?

That’s the end of my questions. Is there anything else you would like to add?

## Data analysis

We utilised an abductive analytic approach,^19^ whereby data collectors initially entered verbatim quotes relating to barriers and solutions into a deductive framework matrix, nesting each quote under one of ten broad *a priori* themes that had emerged from a literature review that is described in our protocol:^31^

- Costs
- Distance and transport
- Desire/priority to seek care
- Clinical service quality
- Facilities
- Awareness & communication
- Fear
- Norms, values, health beliefs
- Empowerment, support & capacity
- Other (making room for surprising/unexpected themes)

At daily debrief sessions, SK and LA reviewed the matrix with the data collectors and used inductive coding to identify unique barriers and solutions. The decision to use an analytic matrix and collective interpretation was based on the findings of our previous systematic review, which had found these techniques to be rapid and robust.^30^

Our matrix had one participant per column and one sub-theme per row – with a new row created every time a sub-theme (a unique barrier or solution) was identified. Each sub-theme (e.g. ‘loss of earnings’) was nested under the relevant theme (e.g. ‘costs’) The process of data entry is demonstrated in this short online video (http://tinyurl.com/29asc6nm) and a blank matrix template is available <u>here</u>.

We generated one matrix for the telephone interviews and another for the in-person interviews. This was so that we could compare the themes that emerged from each modality in our embedded study. For our main analysis, presented here, we pooled all barriers and solutions identified using both modalities.

We trained the data collectors over two days and performed fourteen pilot telephone interviews before starting data collection. Videocall debriefing sessions were held at the end of each day.

### Additional analysis

Our original equity analysis had also indicated that people with the highest incomes and those who owned a car or truck may have been less likely to attend that those reporting no vehicle ownership and lower incomes. We conducted an additional ten interviews with people who reported earnings in the highest income category to assess whether the barriers they reported differed from those reported by younger adults. We hypothesised that richer people did not access VIP services because they had sought private care after being identified with an eye need during eye screening.

### Output and screening

We created a summary list of all of the unique solutions that had emerged from the interviews. Before taking these to a representative sample for ranking, we met with the implementing partner to identify any ideas that would be completely infeasible given the constraints of the programme e.g. providing helicopter transportation. Any interventions that were deemed to be completely infeasible were removed from the list. We asked the director of Peek Vision to independently review these decisions.

### Stage 2: telephone survey

#### Survey instrument

We used the vetted list of solutions to generate a simple telephone-based survey (Appendix 2) where respondents were asked to rank each suggestions from 1-3 on a Likert scale:

1. It would make a big difference - i.e. if we introduced this change then you or people like you would definitely attend,
2. It would make a moderate difference - i.e. it would greatly increase the chances, but it would not be enough to guarantee attendance by itself,
3. It would make a small difference - i.e. it might help a few people, but the impact is likely to be minimal.

The telephone ranking survey was piloted with 26 people.

#### Sampling and recruitment

We used a 95% confidence interval, a 5% margin of error, and a conservative assumption that the total population size was 1 million people, rendering a minimum sample size of 384. We took the same list of 18–44-year-olds who had not been able to access care, and used random numbers to generate a call order, removing those who had already been included in the qualitative interviews. The same six data collectors tried calling each person three times before moving on to the next.

#### Data collection and Analysis

Data collectors obtained recorded informed verbal consent, and then read through the survey instrument using an online data entry form. Data collectors entered the respondents’ score for each proposed solution. We calculated the simple mean for each solution, and then ranked solutions by mean score.

### Stage 3: multistakeholder workshop

Once we had this ranked list of solutions, we convened an online workshop with representatives from the programme implementer, programme funder, the county and national health ministry teams, and our community advisory board. We facilitated a discussion where each stakeholder shared their perceptions of the likely impact, feasibility, costs, and risks associated with each solution. As external public health and research ‘experts’, LA and SK restricted their contributions to presenting the ranked solution scores, facilitating the discussion, and providing information on the general strength of the international research evidence for each of the proposed solutions. At the end of the workshop, we asked the participants to collectively agree on one or more solution to implement in the VIP programme. Figure 1 provides an overview of our entire approach.

**Figure 1:**
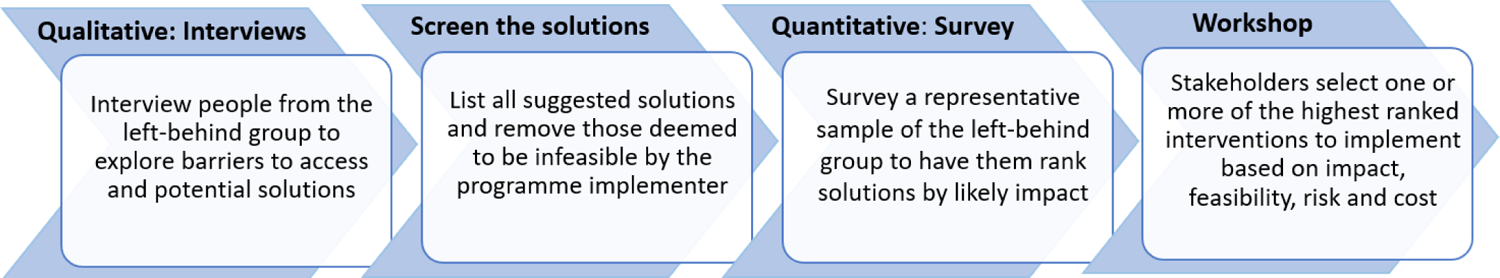
Overview of the sequential mixed-methods approach

This study was approved by KEMRI and LSHTM ethics committees. Those who attended in-person interviews were given a transport reimbursement of KES 500 (USD 3). We used the COREQ checklist to report our study (Appendix 3).

### Findings

#### Interviews

We made 143 phone calls to invite people to participate in in-person and telephone interviews. Three people declined; 29 did not pick up after three calls; and 34 people agreed but either were not home (13 people) or did not arrive at the agreed interview location (21 people) on the day of their in-person interview; six were not eligible as they told us they had actually received care (i.e. they had not been checked-in properly); and four had moved to a different part of the country. In total we conducted 36 telephone interviews and 31 face-to-face interviews over the course of eight days in September 2023. All our participants were aged 18-44 years old and 53.2% were male.

We ended up performing more interviews than were needed to achieve thematic saturation with the 12+2 approach due to the efficiency of our data collectors. At the debrief on day two, they had already completed 24 telephone interviews. Our research leads had not assessed whether saturation had been reached by the time of the call, so – erring on the side of caution - they advised completing a further day of interviews. By the end of day three, 36 telephone interviews had been completed. A detailed retrospective saturation analysis, presented in Appendix 4, concluded that approximately 30 interviews were required to reach thematic saturation (Figure 1). We conducted 31 in-person interviews to enable fair comparison between telephone and in-person interviews for our embedded study.

**Figure 2:**
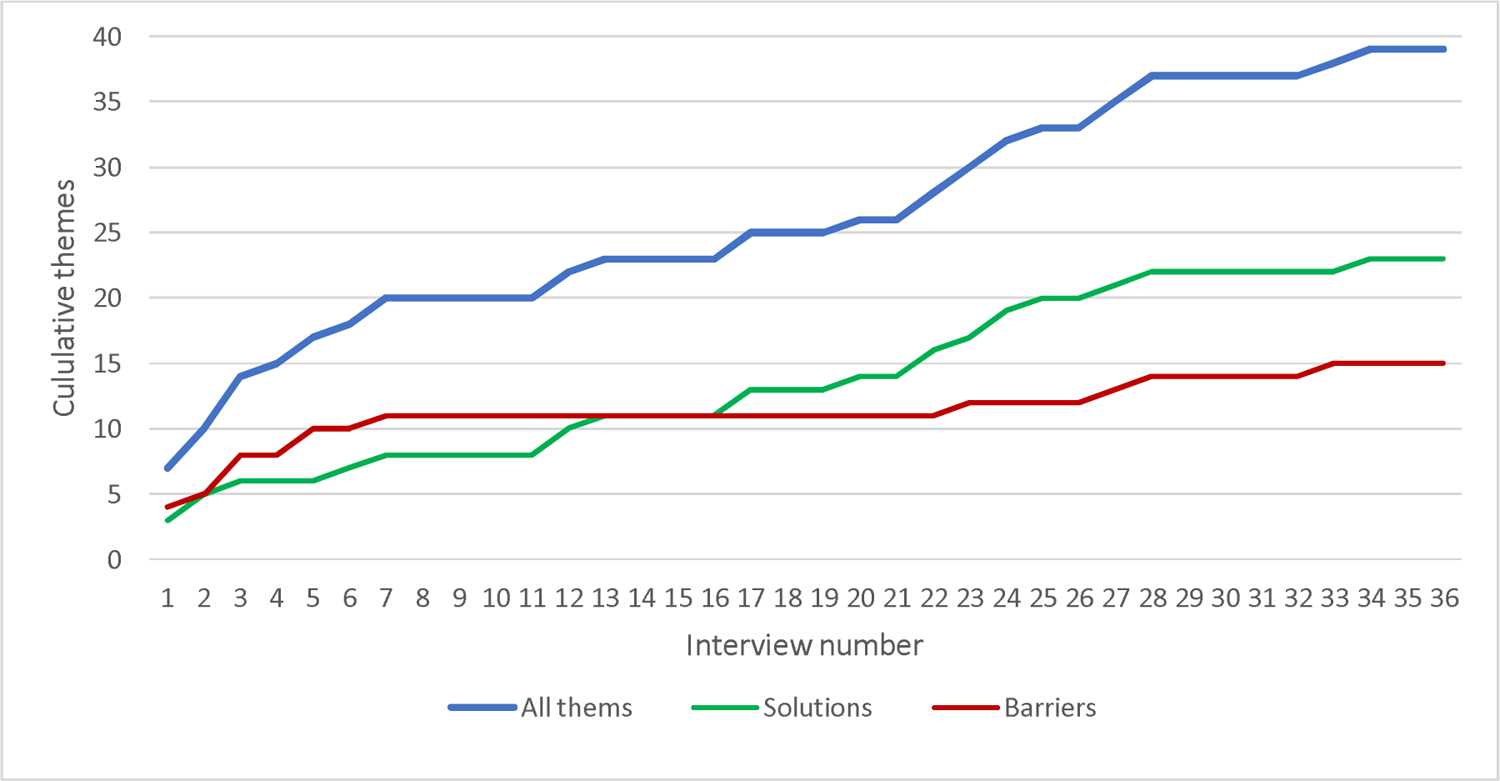
Accumulation of themes as the interviews progressed

Supplementary Table 1 (Appendix 5) presents the 21 unique barriers that were identified along with all the quotes from both sets of interviews. Direct, indirect, and opportunity costs; long queues; difficulty getting time off work; and insufficient information about opening times and dates emerged as important themes. We also identified several meta-themes; participants were generally able to access the clinic locations but left after seeing long queues of several hundred people. Many felt they could not ‘waste time’ waiting to be seen, given the associated loss of potential earnings.

> “I choose to go to work to make money rather than spend my days’ time not knowing whether I will get attended to… If I don’t work, I don’t get money. MFK008, in-person

Another important cross-cutting theme was the perceived lack of information about the clinics: where they were, days of operation, opening and closing times, and what services were available. Assumptions around (non-existent) costs and early closures also prevented some people from attending.

> “I also forgot the exact location where I was to go for the eye check-up and no one followed up to remind me of the place and date.” MT33, telephone

> “They did not tell us if we need to come with money or not. Eye treatment, we are usually told to come with money. I assumed I’ll go there and they will ask me for money and I did not have it.” MFK204, in-person

One interviewee also told us that the counselling he had received at the point of referral was inadequate. He wanted more information about what would happen at the clinic and on why attending was important, especially given that he did not even realise he had a problem:

> “They just told me that I have problems with my eyes and I should visit [town name] dispensary so I did not know what I was going to do there, is it surgery, is it being given medication, is it being tested again? And for me I have always known that my eyes are okay, and on that day they told me that they are not okay. They were very brief and I didn’t know what to expect, so that shock of being told that I have an eye problem which I have never had before is the reason why I did not go.” MFI03, in-person

In terms of novel barriers, one person told us that they left the queue because they were “an introvert” and didn’t like the crowd (MT772, telephone); another felt their eye problem needed emergency treatment and sought care elsewhere (MFK02, in-person). One interviewee specifically named male health seeking behaviour as the main reason he didn’t attend:

> “As a man it is very hard to prioritize my health as I am manly focused on my family’s wellbeing and It is easy to forget my health needs” (MT250Z, telephone).

Finally, one young man explained that being made to queue alongside women and children was shameful:

> “There were women on the line. They could have different lines for youths and women for some us to be comfortable because it is shameful to be on the same line with women and children, with worries how they will perceive me as young man. It was a challenge for me to just stand there with women… I had to go back that day without being attended even though right now my eyes have a problem. MT040, telephone

The 25 proposed solutions to improve access centred around reducing the clinic queue lengths so that people could be seen quickly and then get back to work. Ideas included adding more clinics, holding them closer to villages or workplaces, increasing staff punctuality and speed, scheduling fewer people to attend each day, and extending the opening days and hours. The other meta-theme related to the provision of more detailed information around clinic services and opening times. Table 1 presents a summary of all 25 suggested solutions along with illustrative quotes. A full list all solution-related quotes is presented in Appendix 6.

#### Reviewing feasibility

As planned, we presented the list of all 25 suggestions to senior representatives from the implementing partner. We asked them to identify any suggestions that would be completely infeasible to deliver, given that they are responsible for funding all aspects of the programme. They felt that the programme budget would not permit additional payments for transport reimbursement or attendance incentives. Given that the outreach clinics involve multiple members of staff and large volumes of equipment and supplies, they also felt that it simply wouldn’t be feasible to deliver a door-to-door version of the outreach clinic. These suggestions were removed from the list. The director of Peek Vision agreed with each of these decisions. The remaining 21 suggestions were put to a representative sample of people from the left behind group in a ranking survey.

**Table 1:**
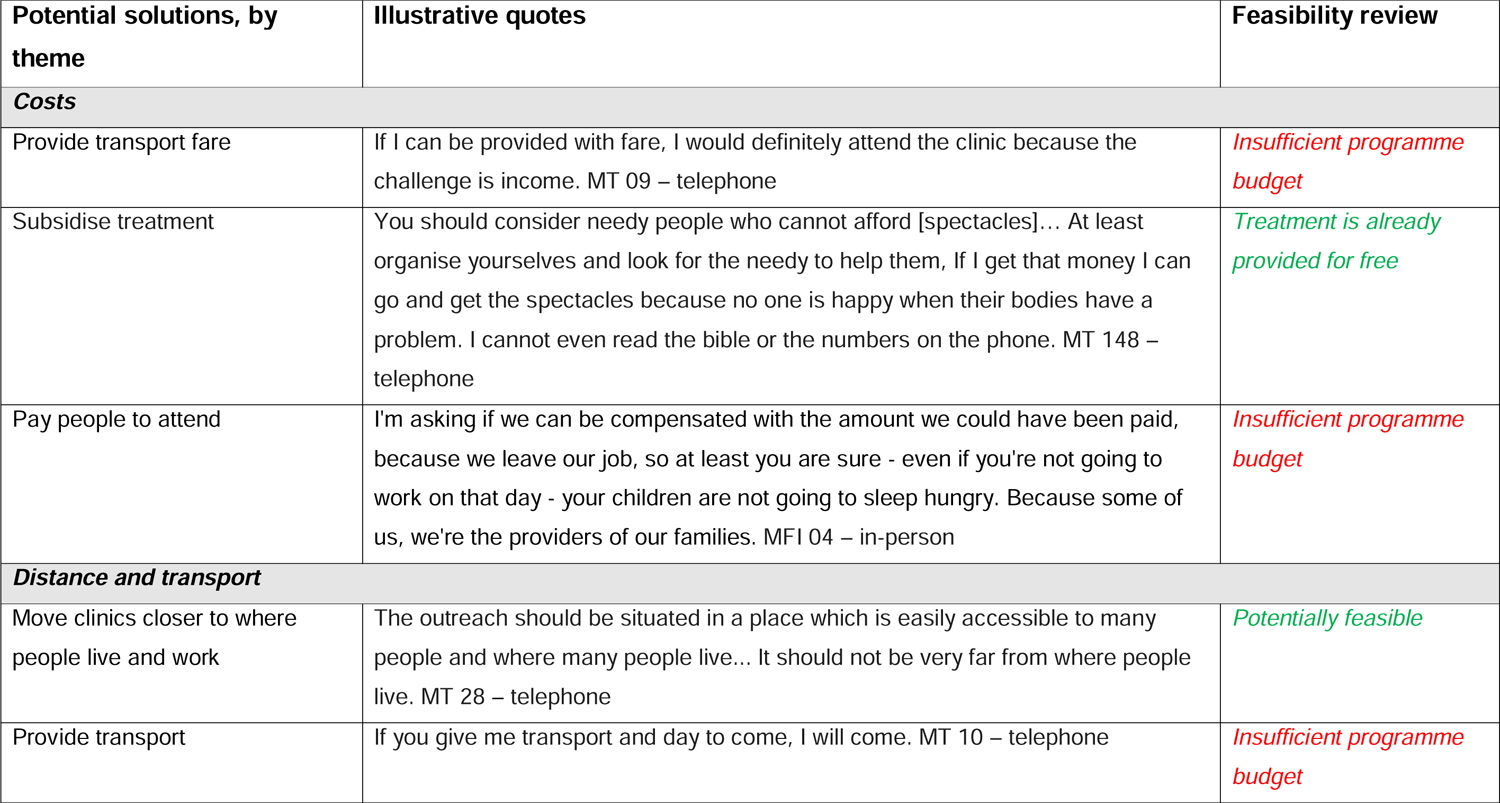
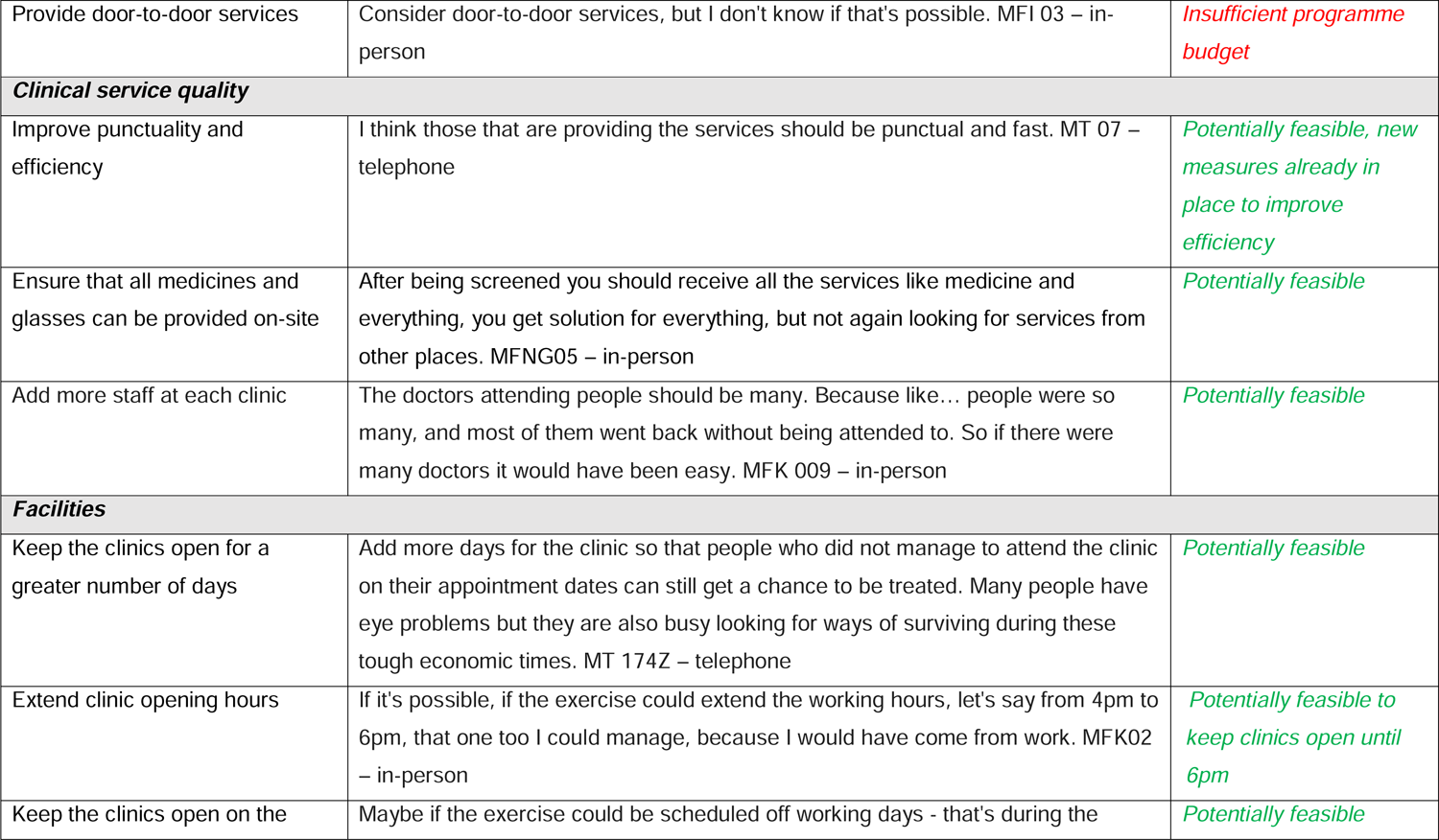
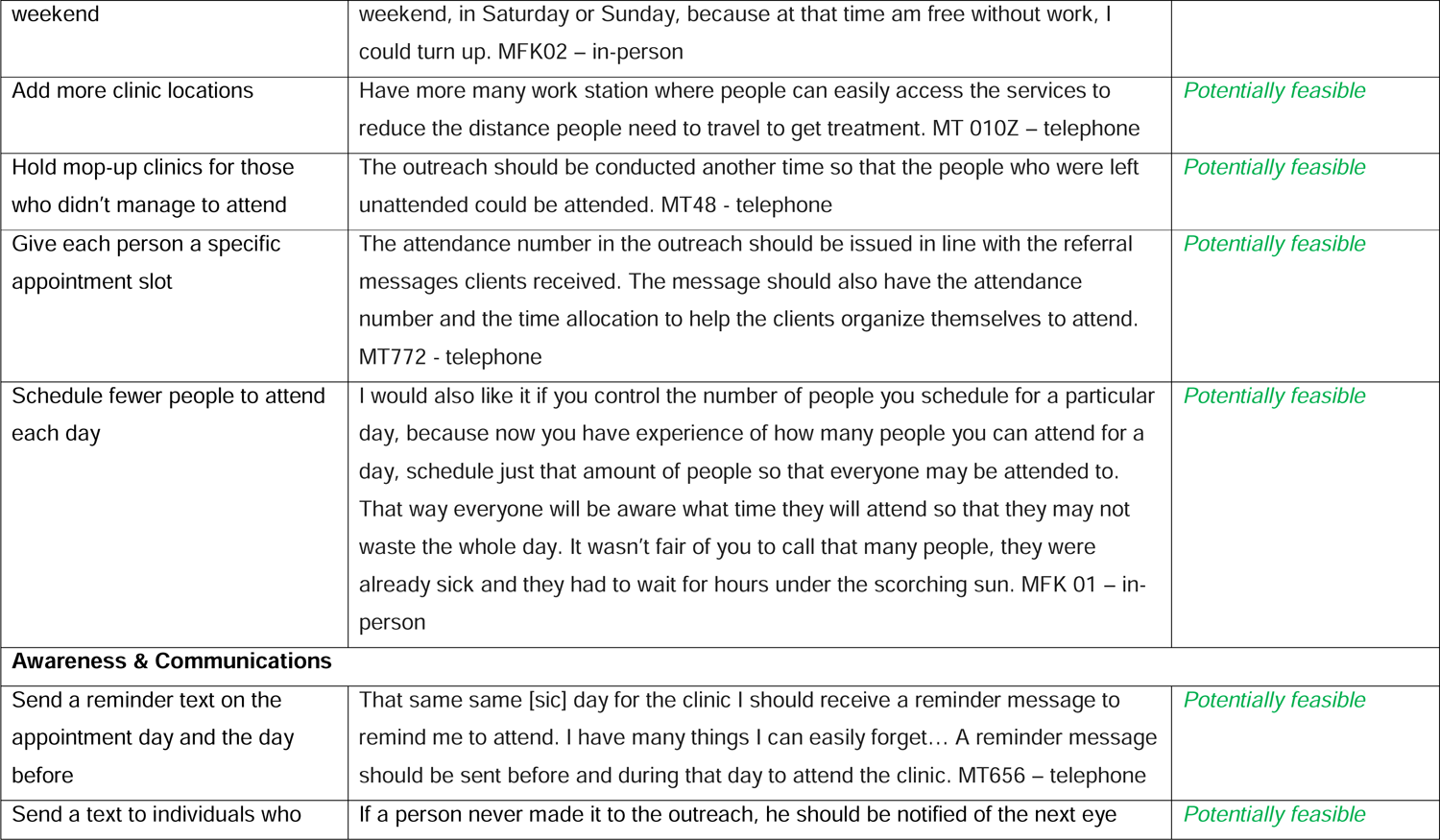
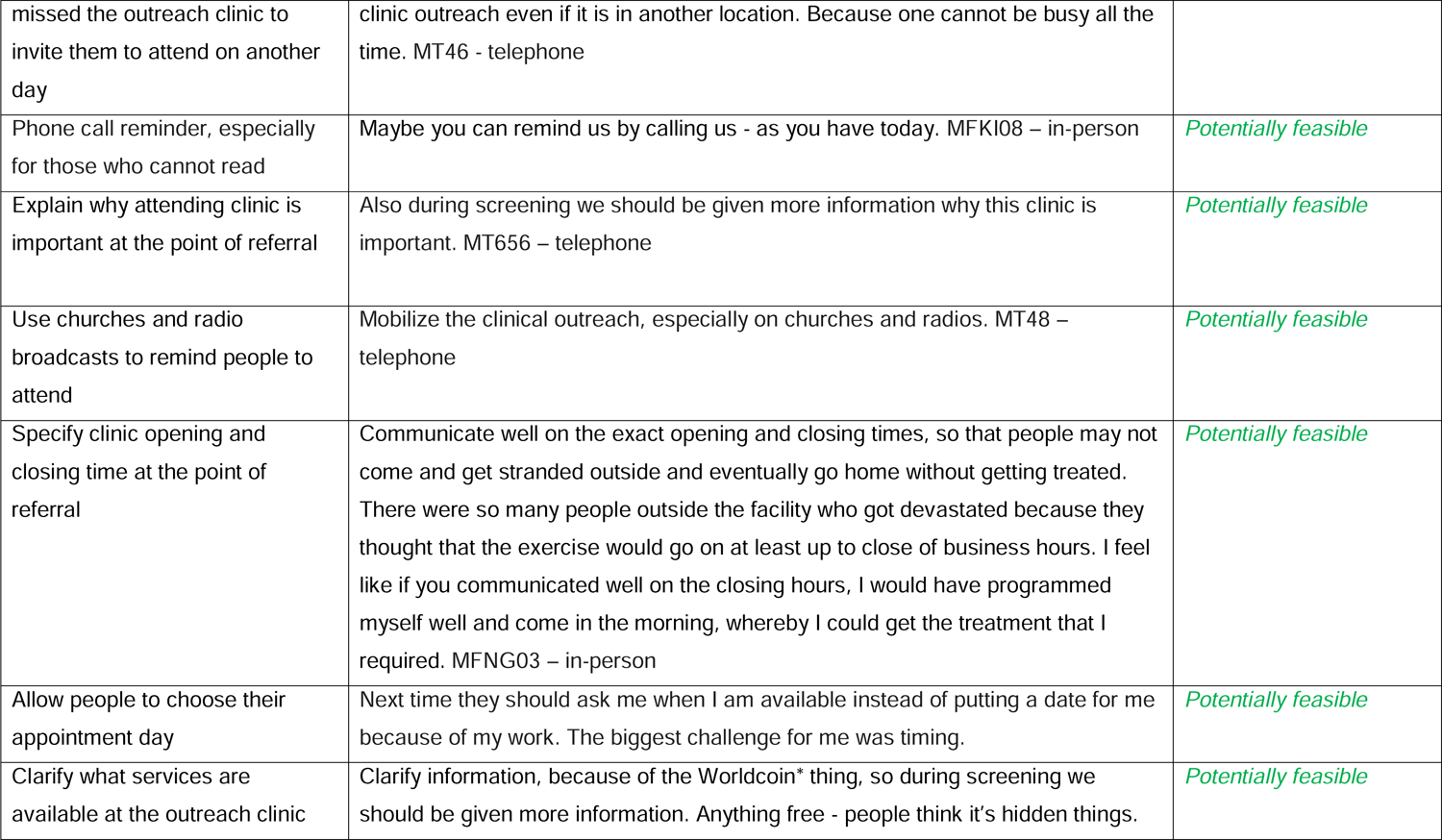
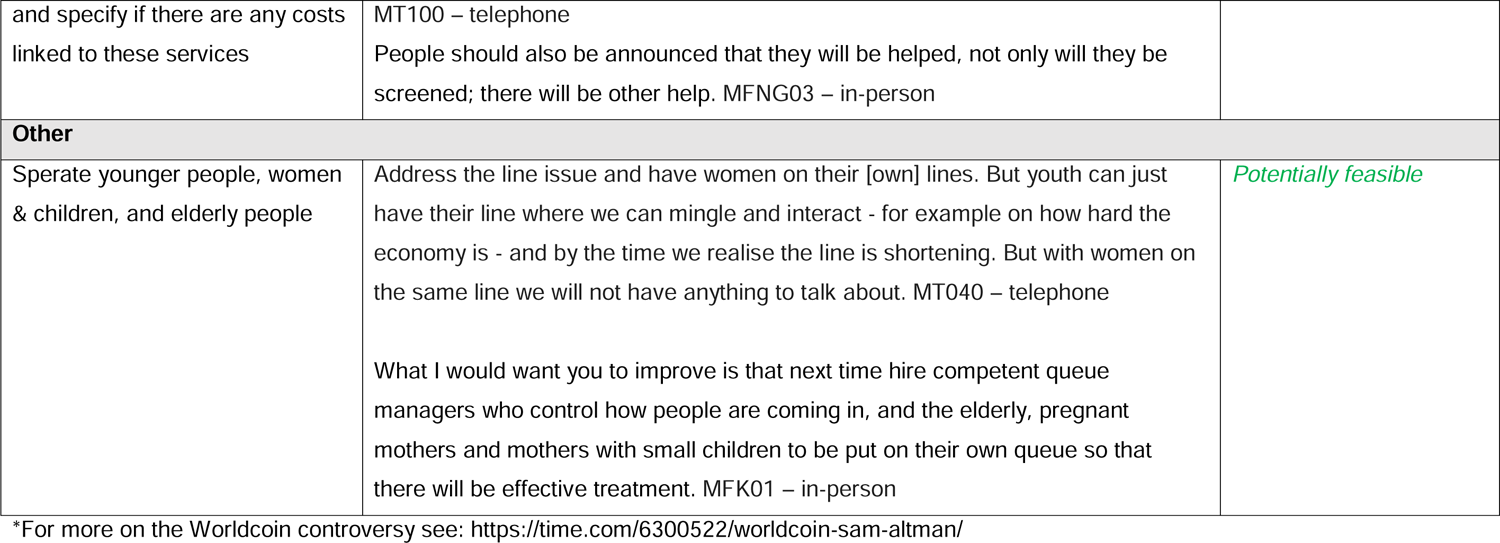
Solutions and implementing partner feasibility assessment.

#### Survey and selection workshop

It took two days to train the data collectors and pilot the survey, and five days to complete ranking exercise. In total, data collectors called 440 people, of whom 401 completed the survey (response rate = 91%).

All of the suggestions received a mean score between 2.4–2.9, indicating that all of the potential solutions were felt to offer at least a moderate chance of improving attendance. Table 2 presents the scores for individual-level solutions (that could be tested in an individually randomised RCT) and solutions that would be implemented at the clinic level (requiring testing with cluster RCTs).

Our online workshop included representatives from the community advisory board, the screening programme implementing partner organisation, the programme funder, the national eye screening programme office, Kenya Society for the Blind, and the county health department. After our team had presented the survey findings, we facilitated a discussion around each of the proposed solutions in turn.

The majority of the suggested clinic-level interventions required additional human resources or clinic locations. The programme funder and implementing partner recognised the issues around long queue times in some locations, but were very clear that inflation had already taken the programme over budget and there were no spare resources to introduce additional clinics or staff. The top-ranked suggestion related to frustration experienced by people who attended the treatment outreach clinic but were found to have a complex eye problem that required onward referral to the local hospital for specialist spectacles (where care is subsidised but not free). The workshop participants also recognised this problem, but agreed that it was not possible to have advanced ophthalmic care services present at each outreach clinic. The group suggested clarifying the process of tiered referrals during counselling. The workshop participants felt that organising separate queues for different ages and genders would be practically feasible, however imposing separation may cause problems for friends/family/colleagues who attend together. After reviewing all of the suggestions, the workshop participants unanimously agreed to implement and test the following bundle of interventions relating to counselling and the provision of enhanced information in SMS reminders:

– Send a reminder text on the appointment day and the day before
– Clarify what services are available at the outreach clinic and costs linked to these services
– Specify clinic opening and closing time at the point of referral
– Explain why attending clinic is important at the point of referral

Overall, the ethics review process took four months. The interviews took nine days to complete, and the survey took seven days, including training and piloting. We held the multistakeholder workshop one month after the survey had concluded, with the delay driven by scheduling challenges.

**Table 2:**
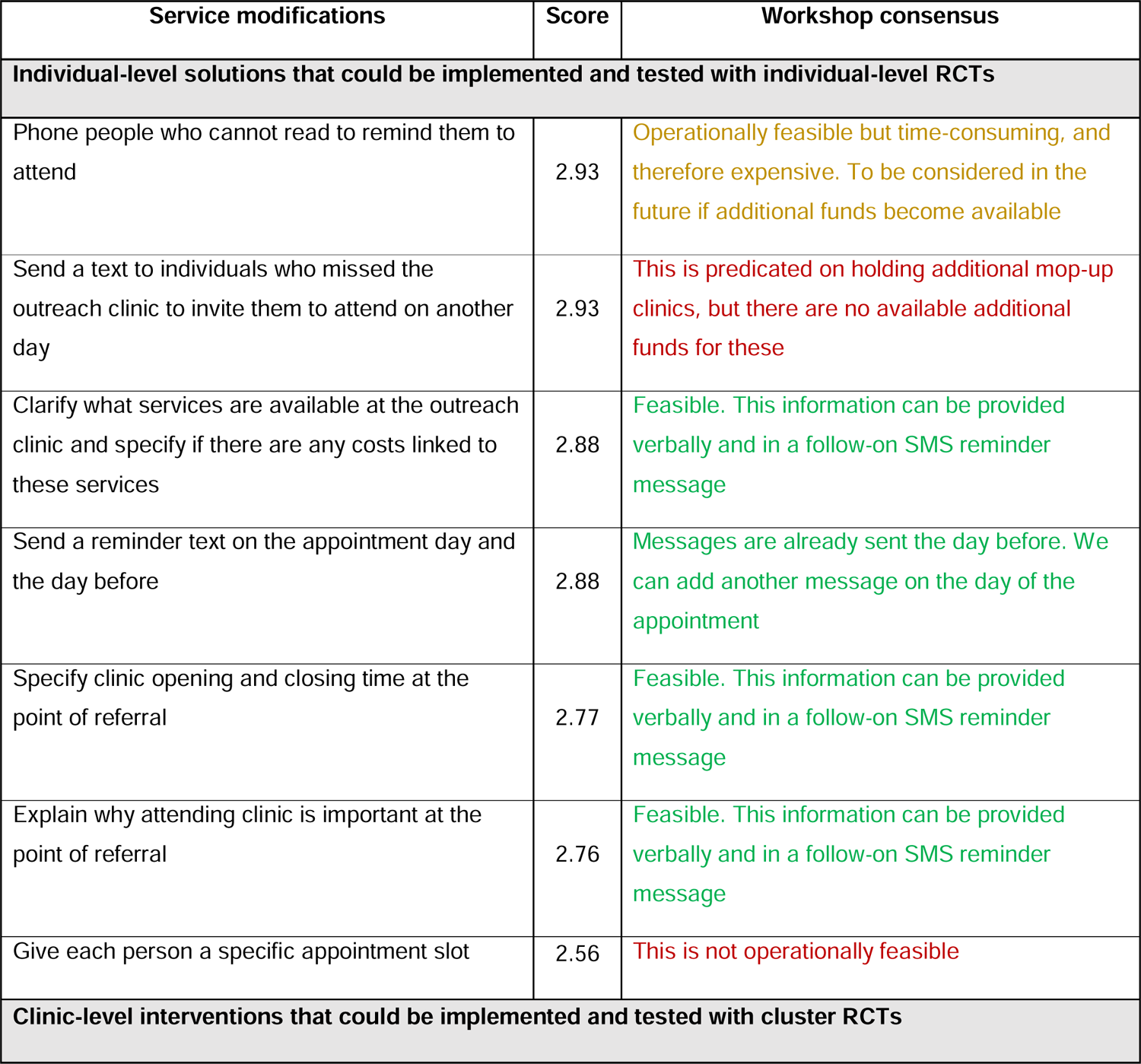
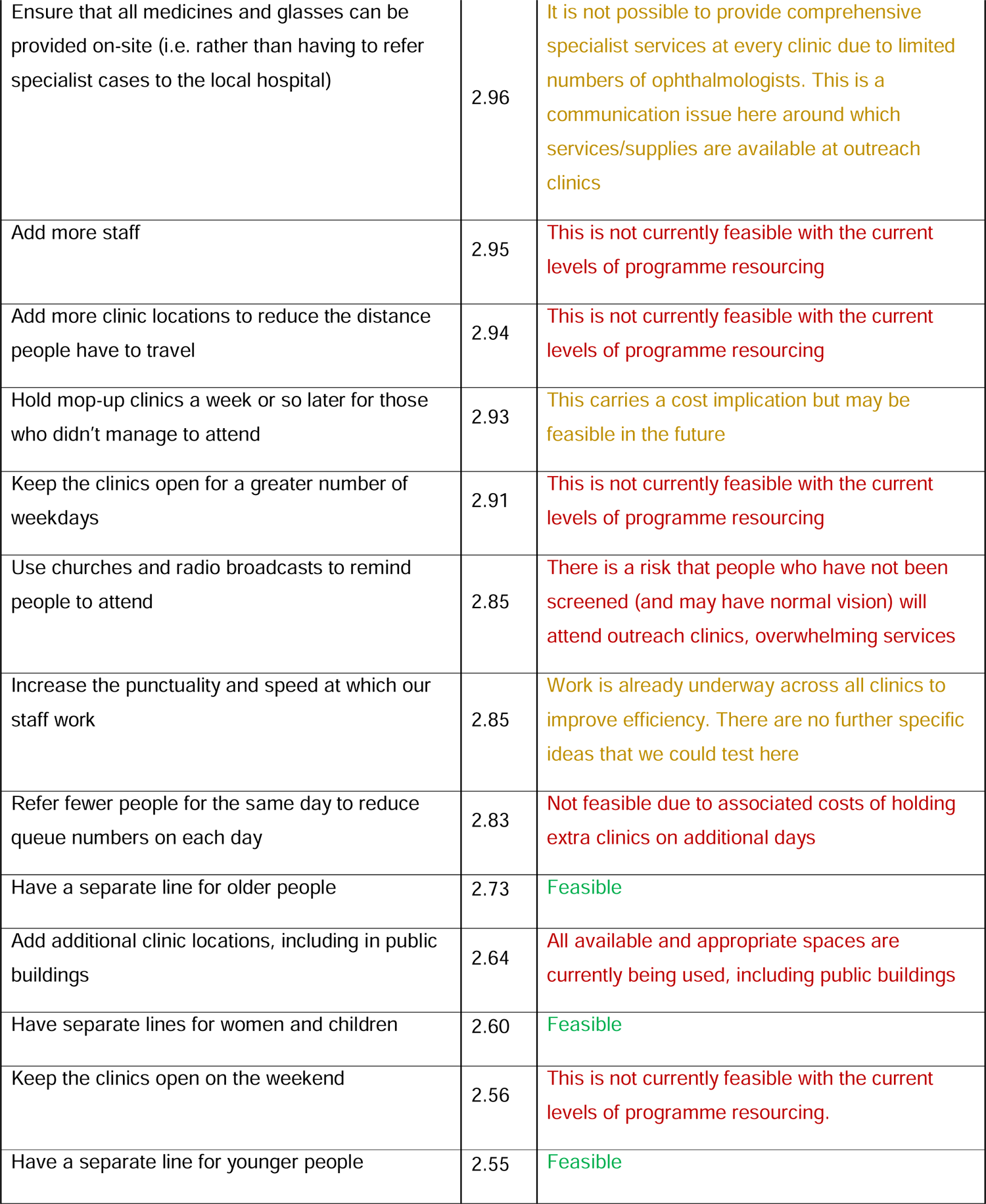
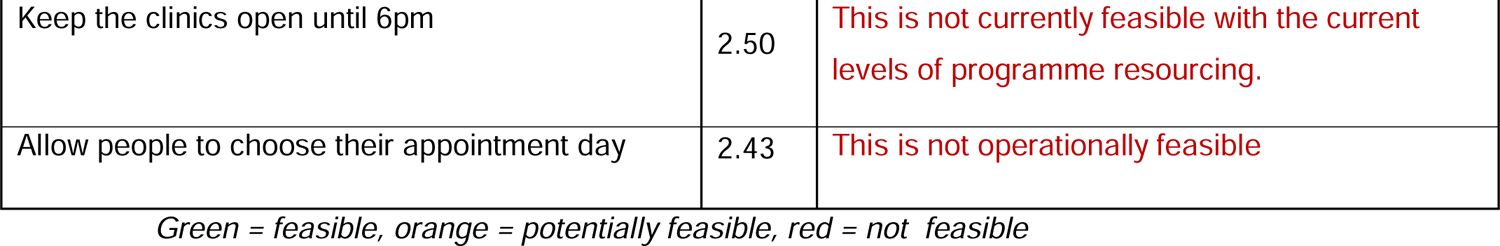
Mean scores for each of the suggested solutions and workshop consensus.

The current verbal counselling script and SMS reminder messages that are used in the VIP programme are presented in Box 2. The SMS reminders are currently sent on the day of referral and the day before the clinic appointment. We drafted a new verbal counselling script and SMS reminder message that included the suggested new elements that were agreed in the workshop. We asked the workshop participants to review the new wording via email, as well as two representatives from the left-behind group. Based on this feedback we made three minor changes. The original script and description of these changes is presented in Appendix 7. The enhanced counselling and SMS reminder will be tested in an individual-level RCT in a subsequent study.

##### Box 2

###### Original and new counselling and SMS reminder wording

Usual care counselling, delivered verbally at the point of referral

“I have examined your eyes, and you have a problem. I have referred you in the system and you will receive an SMS with where and when you are supposed to attend treatment. You will come for treatment on <<date>> at <<location>>. The examination will be free and you will be informed of anything else on the material day.

Current SMS reminder, sent on the day of referral and day before the appointment

Dear <<name>>, you were examined and found to have an eye problem. Kindly report on <location> on <<date>> for assessment. For more information contact Meru Referral Hospital.

Enhanced counselling script, based on interview, survey, and workshop feedback

“I have found a problem with your eyes. I am referring you to the outreach treatment clinic that will be held at <<location>> on <<date>> between <<time>> and <<time>>. At the clinic, eye care professionals will perform a specialist assessment and provide any eye drops or medicines that you might need. If you need glasses, the specialists will tell you what kind you need, and what your prescription is. The assessment is completely free. Note that a small proportion of people will be found to have complex eye problems that require onward referral for hospital assessment and special lenses that cost more than standard glasses. However, the vast majority of people have their needs fully met at the outreach triage clinic and do not need hospital referral.

With treatment, you will be able to see more clearly. This will help with your work, seeing faces, and using your phone. It is important that you attend the clinic to protect your vision. The clinic will only be running from <<day>> to <<day>>, so if you don’t manage to attend, you may not be able to get free care again in the future.”

Enhanced SMS reminder, based on interview, survey, and workshop feedback, to be sent on the day of referral, the day before the appointment, and on the day of the appointment

We found that you had an eye problem. Please attend the outreach clinic at <<location>> on <<date>> between <<9AM-2pm>>. The specialist assessment is free

If you are found to have a complex problem, you may be referred to a hospital for further care or specialist glasses, and this may include a fee

However, the vast majority of people who attend the outreach get their eye problem fixed without the need for any further referral

It’s important that you attend to protect your vision, and you might not have a future opportunity to access free care. See you on <<date>>

#### Sensitivity analysis

We conducted ten additional interviews with high-income individuals aged 18-44 who had not managed to access care, based on previous evidence that this group were potentially facing unique barriers. We triangulated the themes with those from the other interviews. High-income interviewees did not mention loss of wages as a potential barrier, but the cost of treatment and transport were raised, along with conflicting engagements, long queues, and lack of clear information. No unique or discordant barriers were identified. Similarly, the solutions raised were closely aligned with those from the other interviews, including requests for financial subsidies and incentives – suggesting that this group were not necessarily very affluent i.e. our original ‘high-income’ threshold (USD 2,600/year, aligned with the national top income tax threshold) was set too low. The only unique solutions related to a request for more stylish spectacle frames; “Next time come with spectacles of good standard, not for the old people.” (HIG3); the use of Community Health Promoters to remind people to attend; and asking for permission from employers to get time off. Full quotes and codes are presented in Appendix 8.

## Discussion

We conducted 67 interviews and 401 solution ranking exercises with people from the sociodemographic group that had previously been found to face the greatest barriers to accessing eye care services in Meru – young adults aged 18-44 years. These people told us that lack of clear information, long queues, and the opportunity costs associated with long queues were the main barriers to receiving care. Whilst lack of clear information and inadequate counselling have emerged from multiple previous studies in diverse settings and populations,^32–36^ the theme of long queues does not commonly appear in the wider literature or appear as one of the drop-down barrier options that is used in *Rapid Assessment of Avoidable Blindness* (RAAB) surveys that have been deployed in over 80 countries.^37^ We postulate that the ubiquity and fundamental intractability of this problem in resource-scarce settings leads to them being perceived as the status quo rather than a specific barrier.

Research from several other African countries suggests that most patients generally wait 1-4 hours to receive ambulatory care,^38–43^ and even in a well-resourced setting like the UK, 10% of patients are currently waiting more than 12 hours to receive emergency care.^44^ Younger adults may be more sensitive to long wait times as they are more likely to be in active employment than older adults, and may be more likely to be ‘hustling’ with multiple informal jobs that do not provide protected sick leave, meaning that they experience the greatest potential opportunity costs from waiting in line for several hours.^45^

Rapidly surveying a representative sample of younger adults enabled us to move from subjective to generalisable themes. For instance, if we had stopped after completing the interviews, we would not know whether the issue of mixing men and women in the queues was a major structural barrier to access for this group, an unusual individual quirk, or something in-between. In the event, we found that scores accorded to all of the 21 potential solutions clustered between 2.5-3.0, indicating that members from the left behind group felt that all of the suggestions were moderately-to-highly likely to improve access to care. We cannot exclude the possibility that the universally high scores are at least partially driven by cultural norms and/or a form social desirability bias.^46,47^ We took all of the ranked suggestions to a multistakeholder meeting that included representatives from the left behind group, the local community, the programme funder and implementer, and a national eye charity. This group weighed the rankings against considerations of the risks posed by each intervention, their collective estimation of likely impact, the associated costs, and the operational feasibility of implementing each solution. This group reached a consensus agreement to focus on a bundle of solutions aimed at providing timely and clear information about the outreach clinics.

The selected interventions focus on empowering individuals with the information they need. However, a different perspective is that the workshop participants dismissed more highly-ranked service-side solutions in lieu of minor interventions that place responsibility for access on intended service users, in line with the widely debunked ‘information deficit model’.^48^ Allen has previously argued that efforts to equitably advance UHC and ‘health for all’ should focus on unjust social structures and resist focussing wholly on ‘downstream’ technocratic solutions. In the context of this study, the paucity of good jobs, the absence of adequate social welfare systems and strong labour laws that guarantee paid sick leave are examples of major structural challenges for younger adults, however these were not raised by the interviewees.

The greatest strengths and limitations of our approach pertain to voice and epistemic justice.^13^ We strove to centre our approach around the perspectives of those with lived experience of not being able to access care. The ‘pose’ and ‘gaze’ of the project^15^ focus on the credibility and value of this group, as well as locally recruited data collectors’ work in sensemaking through their initial interpretative work as they engaged with the analytic matrix. Findings from this process fed directly into the local programme, with their implications wholly determined by local stakeholders. Although Europeans and Kenyan public health experts were involved in leading the project, the primary interpretation was locally owned and the resulting solutions will be locally implemented for the benefit of local Meru residents.

The programme funder and implementing partner held veto rights as they were ultimately responsible for delivering service modifications and could not be ‘made’ to implement any of the suggested service modifications. However, community, charity, and public health stakeholders reached one accord with the programme leads on the infeasibility of introducing additional staff of clinics given their shared appreciation of how inflation had literally decimated the programme budget. Nevertheless, our approach involved (and necessitated) epistemic tension; even though all of the solutions came from the left-behind group, the final decision on how to modify the programme required consensus between different stakeholder groups with imperfectly overlapping interests and unequal power in the final determination. We feel that including programme managers is a design strength rather than a flaw, as there is no value in presenting programmes with a *post-hoc* shortlist of unworkable suggestions. Furthermore, the process of bringing them into conversation with community members, eye care advocates, and representatives from the left behind group can create shared understanding and constructive dialogue. Our role as ‘external’ research leads primarily involved developing rapid and robust methods that would surface non-tokenistic and reliable suggestions, as well as convening actors and facilitating the overall process. We aim to work with local partners to embed this approach so that future iterations can be wholly led by local teams.

Finally, our unplanned sampling deviation led us to conduct more interviews than expected. The post-hoc sensitivity analysis presented in Appendix 3 suggests that if we had adherence to our original ‘12+2’ approach, we would have missed four barriers and 12 additional solutions. We agree with Guest and colleagues that current approaches to assessing and reporting saturation in qualitative research are opaque, however our empirical analysis suggests that – for studies like ours – saturation is only reached after three interviews in a row reveal no new themes, after a minimum of 15 initial interviews have already been conducted (the ‘15+3’ approach).^23^

## Conclusions

Interviews and ranking surveys conducted with people who were not able to access eye care services identified 21 barriers and 25 potential solutions, centring around reducing the time it takes to be seen, and providing clearer information about the eye clinics. Multistakeholder workshop participants ended up selecting interventions that did not fundamentally alter how clinics are operated, instead opting for amendments to the way that information is provided. Even so, these service modifications came directly from those who were being left behind, and were rated likely moderately-to-highly likely to improve access. Including service funders and managers in interpretive and deliberative processes means that our findings will result in concrete, locally-led service modifications. Further work will evaluate the impact of these changes using an embedded, pragmatic RCT.

## Data Availability

All data produced in the present work are contained in the manuscript.

## Acknowledgements

We would like to thank Paul Ringera for reviewing the study protocol and providing invaluable community perspectives and input on this project.

## Appendix 1: Call scripts

### Telephone interview consent script and topic guide

Hello, my name is. I am a researcher from KEMRI, working with the Ministriy of Health and the VIP eye screening programme.

Your recently had your eyes screened and were found to need further assessment. Our records indicate that, like many other people, you were unable to attend that appointment.

You are being contacted because you have previously provided consent to be contacted regarding research being conducted for eye care services. I am calling to invite you to participate in a 15-minute interview. Your participation is completely voluntary. This means that you do not have to do it unless you want to.

We want to understand the barriers that prevented you from attending. We are also asking about how we could change the VIP programme to make it easier for people like you to attend appointments.

Before agreeing, here is the background information that you need to know:

We have invited you because, like many other people, you did not attend. We want to hear about the issues that you personally faced that prevented you from attending, and your ideas on how to make things easier. In total we are aiming to interview about 20 people.

Who are we? I work with a group of researchers from the KEMRI and the London School of Hygiene and Tropical Medicine. We are working to improve the VIP eye screening programme. The leaders of the research are Prof Michael Gichangi and Dr Luke Allen.

We will take the responses from all of the interviews and discuss the ideas for improvement with the leaders of the national programme. We hope to use your suggestions to make the programme work better.

We are also conducting a set of face-to-face interviews with other people who did not manage to access care. We want to compare the responses we get from these different approaches.

In this 15-minute interview there are no risks or benefits to you. It is important to note that agreeing or declining to take part does not have any impact on the services you receive.

You can stop the interview at any time.

I will record the interview. Our team will anonymise your data and keep it safe and secure on a password-protected computer in London. When the study is completed, we will write-up our findings and publish them online so that other researchers can use the information to help people in other places.

The KEMRI and London School of Hygiene and Tropical Medicine ethics committees have both approved this study.

You can ask me any questions you like now. I can also give you the email address and phone number of the lead researchers if you’d like to contact them directly [provide the contact details for Sarah Karanja or Luke Allen as required]. If you have any other concerns I can also give you the contact details for the London School of Hygiene and Tropical Medicine Research Governance and Integrity Office.

Do you have any questions?

Are you happy to begin the interview?

[If yes, proceed; if no thank them for their time and end the call]

### Opening questions

To start with, during the home visit by the vision impact project team, please describe your experience with the eye screening.

Probes:

- How long did the examination take?
- Was the test comfortable? If yes, why? If no, why not?

### Barrier elicitation questions

You were assessed at home and found to have an eye problem that required further assessment. You were asked to attend the community eye clinic on [date] and reminders were sent to this mobile number.

- In your own words, can you talk me through why we didn’t see you at that clinic?

### Probing questions

- Are there any other factors that prevented you from attending?
- Is there anything else you’d like to share?
- Of the issues you mentioned, which is the most important?

### Solution elicitation questions

The last part of the interview is exploring whether there is anything we could do to address these barriers and make it more likely that you will attend in the future.

- So to start, what would make the biggest difference?

### Probing questions

- What else would help?
- What other changes could we make to the programme that would make it easier for you to attend?
- Are there any other specific changes that we could make to the way that the programme or eye clinics run?
- You mentioned [list their proposed solutions]. Some of these may be beyond our control, but if we managed to [list their proposed programme-related changes], do you think that would be enough?

That’s the end of my questions. Is there anything else you would like to add?

### Probing questions

- After missing the outreach clinic appointment, did you seek medical care from a different healthcare facility for your eye condition?
- If no, why not?
- If yes, where did you seek medical care?

Thank you so much for your time.

### Call script to invite people to participate in an in-person interview

Hello, my name is. I am a researcher from the Kenya Medical Research Institute (KEMRI), working with the Ministries of Health and Education on the Vision Impact Project eye screening programme.

You recently had your eyes screened at the community outreach programme and was found to need further assessment. Our records indicate that, like many other people, you were unable to attend that appointment.

You are being contacted because you have previously provided consent to be contacted by Ministry of Health partner organisations regarding research being conducted for eye care services. I am calling to invite you to participate in an in-person interview in the next few weeks. Your participation is completely voluntary. This means that you do not have to do it unless you want to.

If you agree, I will arrange to meet you in or near where you live to ask you some questions about the barriers that prevented your child from attending. I also want to ask about how we could change the Vision Impact Project to make it easier for people to attend appointments.

To compensate you for your time you will receive a [KES 500 equivalent] airtime voucher.

If you are interested, I can send you the full study information via WhatsApp or email or talk it through on the phone with you now.

[switch to phone PIL script here as required]

## Appendix 2: Quantitative Ranking Survey

Interviewer name

Short-answer text

Participant ID

Short-answer text

Consent

Good morning/afternoon

My name is and I’m calling from the Vision Impact Project eye screening programme. We saw you a few weeks ago and referred you to the local clinic, but we did not see you on your appointed day.

In fact, half of all people who were referred did not attend. We have sought feedback on ways we could improve our service, and I wanted to ask you which of the ideas we have stand the best chance of helping people like you to access care. It should take approximately 15 minutes of your time.

If you are happy to proceed, I need to tell you a bit more about the survey. I will then double-check that you are still happy to proceed.

I will ask you about a set of potential changes that we are thinking about making. I will ask you to rate each one in terms of how likely you think it is to make a difference at helping people access our clinics.

Your responses will help us to shape and improve our services for others, but there are no direct benefits to you for taking part. Thinking about the issues that prevented you from getting care may be distressing to you. If you face any discomfort because of the questions asked, you can skip any question or ask to end the call whenever you choose.

If you don’t want to take part, that’s ok. You can drop out of the survey at any point. Your decision will not affect your health care or your future relations with the Vision Impact Project in any way.

Your anonymised answers will be combined with those from other people and kept safe and secure on password-protected computers in Nairobi and London. None of the data will be used for commercial use. We will publish our findings in a research journal and in a public repository so that other researchers can learn from what we find. You personal information will not be included in our findings and there is no way that you can be identified from any of the reports that we will produce.

If you have any questions, you can ask me now, or I can put you in contact with the study coordinator - Sarah Karanja from Kenya Medical Research Centre. If you have any questions about your rights as a research participant, I can connect you with the Kenya Medical Research Centre Ethics team who approved this survey.

Does that all make sense? Do you have any questions for me? Are you happy for me to start?

The first two potential improvements are aimed at making it easier for people to get to a clinic.

There are three options to choose from:

1. It would make a small difference - i.e. it might help a few people, but the impact is likely to be minimal
2. It would make a moderate difference - i.e. it would greatly increase the chances, but it would not be enough by itself to guarantee attendance by itself
3. It would make a big difference - i.e. if we introduced this change then you or people like you would definitely attend

If we add more clinic locations to reduce the distance people have to travel, how likely is that to make a difference?

1 []
2 []
3 []

Expand the list of referral clinics to include additional public health facilities and private and faith-based hospitals

1 []
2 []
3 []

The next set of improvements are about reminding people about the clinic

Explain why attending clinic is important at the point of referral

1 []
2 []
3 []

Clarify what services are available at the outreach clinic and specify if there are any costs linked to these services

1 []
2 []
3 []

Send a reminder text on the appointment day and the day before

1 []
2 []
3 []

Send a text to individuals who missed the outreach clinic to invite them to attend on another day

1 []
2 []
3 []

Phone people who cannot read to remind them to attend

1 []
2 []
3 []

Use churches and radio broadcasts to remind people to attend

1 []
2 []
3 []

The next set of improvements are about extending clinic opening hours

Keep the clinics open for a greater number of weekdays

1 []
2 []
3 []

Specify clinic opening and closing time at the point of referral

1 []
2 []
3 []

Keep the clinics open until 6pm

1 []
2 []
3 []

Keep the clinics open on the weekend

1 []
2 []
3 []

Hold mop-up clinics a week or so later for those who didn’t manage to attend

1 []
2 []
3 []

Allow people to choose their appointment day

1 []
2 []
3 []

The next set of improvements are about reducing waiting times

Add more staff to reduce the queue waiting time

1 []
2 []
3 []

Refer fewer people for the same day to reduce queue numbers on each day

1 []
2 []
3 []

Give each person a specific appointment slot

1 []
2 []
3 []

Increase the punctuality and speed at which our staff work

1 []
2 []
3 []

Have a separate line for younger people

1 []
2 []
3 []

Have separate lines for women and children

1 []
2 []
3 []

Have a separate line for older people

1 []
2 []
3 []

The final set of improvements are about stocks & supplies

Ensure that clinics are fully stocked so that all medicines and glasses can be provided on-site

1 []
2 []
3 []

Thank you so much for your time, we really appreciate your generosity, your responses will help us to improve the service Is there anything else you would like to feed-back to us?

Long-answer text

Thank you again, goodbye

## Appendix 3: COREQ checklist

| Topic | Item No. | Guide Questions/Description | Reported on Page No. |
| --- | --- | --- | --- |
| **Domain 1: Research team and reflexivity** |  |  |  |
| *Personal characteristics* |  |  |  |
| Interviewer/facilitator | 1 | Which author/s conducted the interview or focus group? | 9, 10 |
| Credentials | 2 | What were the researcher's credentials? E.g. PhD, MD | 1, 2, 3 |
| Occupation | 3 | What was their occupation at the time of the study? | 1, 2, 3 |
| Gender | 4 | Was the researcher male or female? | 9, 10 |
| Experience and training | 5 | What experience or training did the researcher have? | 1-3, 9, 10, 11 |
| *Relationship with participants* |  |  |  |
| Relationship established | 6 | Was a relationship established prior to study commencement? | 9, 36, 38 |
| Participant knowledge of the interviewer | 7 | What did the participants know about the researcher? e.g. personal goals, reasons for doing the research | 36, 38, 39 |
| Interviewer characteristics | 8 | What characteristics were reported about the interviewer/facilitator? e.g. Bias, assumptions, reasons and interests in the research topic | 9 |
| **Domain 2: Study design** |  |  |  |
| *Theoretical framework* |  |  |  |
| Methodological orientation and Theory | 9 | What methodological orientation was stated to underpin the study? e.g. grounded theory, discourse analysis, ethnography, phenomenology, content analysis | 8, 9, 11 |
| *Participant selection* |  |  |  |
| Sampling | 10 | How were participants selected? e.g. purposive, convenience, consecutive, snowball | 8, 9 |
| Method of approach | 11 | How were participants approached? e.g. face-to-face, telephone, mail, email | 9 |
| Sample size | 12 | How many participants were in the study? | 14, 23 |
| Non-participation | 13 | How many people refused to participate or dropped out? Reasons? | 14, 23 |
| *Setting* |  |  |  |
| Setting of data collection | 14 | Where was the data collected? e.g. home, clinic, workplace | 10, 12, 13 |
| Presence of non-participants | 15 | Was anyone else present besides the participants and researchers? | 10, 13 |
| Description of sample | 16 | What are the important characteristics of the sample? e.g. demographic data, date | 14 |
| *Data collection* |  |  |  |
| Interview guide | 17 | Were questions, prompts, guides provided by the authors? Was it pilot tested? | 10-12, 22-23, 3 |
| Repeat interviews | 18 | Were repeat interviews carried out? If yes, how many? | N/A |
| Audio/visual recording | 19 | Did the research use audio or visual recording to collect the data? | 9, 10, 13 |
| Field notes | 20 | Were field notes made during and/or after the interview or focus group? | 10, 11 |
| Duration | 21 | What was the duration of the interviews or focus group? | 36, 40 |
| Data saturation | 22 | Was data saturation discussed? | 9, 14, 29, 47-54 |
| Transcripts returned | 23 | Were transcripts returned to participants for comment and/or correction? | 10 |
| **Domain 3: analysis and findings** |  |  |  |
| *Data analysis* |  |  |  |
| Number of data coders | 24 | How many data coders coded the data? | 10, 11 |
| Description of the coding tree | 25 | Did authors provide a description of the coding tree? | 11 |
| Derivation of themes | 26 | Were themes identified in advance or derived from the data? | 11 |
| Software | 27 | What software, if applicable, was used to manage the data? | 11 |
| Participant checking | 28 | Did participants provide feedback on the findings? | 12, 13, 17 |
| *Reporting* |  |  |  |
| Quotations presented | 29 | Were participant quotations presented to illustrate the themes/findings?<br>Was each quotation identified? e.g. participant number | 10, 11, 55-83 |
| Data and findings consistent | 30 | Was there consistency between the data presented and the findings? | 14-17 |
| Clarity of major themes | 31 | Were major themes clearly presented in the findings? | 18-22 |
| Clarity of minor themes | 32 | Is there a description of diverse cases or discussion of minor themes? | 12, 27 |

Developed from: Tong A, Sainsbury P, Craig J. Consolidated criteria for reporting qualitative research (COREQ): a 32-item checklist for interviews and focus groups. *International Journal for Quality in Health Care*. 2007. Volume 19, Number 6: pp. 349 – 357
**Once you have completed this checklist, please save a copy and upload it as part of your submission. DO NOT include this checklist as part of the main manuscript document. It must be uploaded as a separate file.**

## Appendix 4: Post-hoc saturation analysis

We originally set out to use Guest and colleague’s simple formula for assessing whether thematic saturation had been achieved.[1] They describe saturation as “the point in data collection and analysis when new incoming data produces little or no new information to address the research question”, and identify three components that can be used to establish when this point occurs:

– The base, i.e. the minimum number of interviews to be conducted before calculating whether saturation has been achieved. It is similar to the ‘initial analysis sample’ described by Francis et al.[2] Guest et al suggest base sizes of 4-6, based on a review of the literature.
– The run length, i.e. the number of interviews that we review to assess whether any new themes have emerged, with reference to those that have already emerged in the base. Guest et al suggest conducting runs of 2-3.
– The new information threshold, i.e. the number of new themes identified in each run as a proportion of the themes identified in the base interviews. Guest et al suggest that researchers may want to stipulate that saturation is only achieved when a run identifies no new themes (new information threshold = 0%), or that the new themes identified represent ≤5% of the themes identified in the base interviews.

We set out to use a conservatively large base size of 12 interviews, based on a literature review presented in our protocol, with runs of two, and a 0% new information threshold. We also decided that we would compare the new information (number of themes) identified in the runs against all previous interviews, rather than just those in the base.

As detailed in our findings section, we ended up conducting 36 interviews. By reviewing the number of themes that were identified in each interview, we are able to retrospectively assess the point at which saturation would have been reached using a range of different base, run, and new information threshold permutations.

For our initial analysis (Table 1), we used a base of 12 and compared new information obtained from each run of two interviews against the themes identified from the original base of 12 interviews. We performed individual analyses for barriers and solutions. We highlighted the points at which the new of 0% was met.

**Table 1:** Saturation for barriers and solutions with runs of two compared to a base of 12.

| Interview number | Barriers |  |  |  | Solutions |  |  |  |
| --- | --- | --- | --- | --- | --- | --- | --- | --- |
|  | Barriers identified | Novel barriers | Cumulative barriers | New info: runs of 2 | Solutions identified | Novel solutions | Cumulative solutions | New info: runs of 2 |
| 1 | 4 | 4 | 4 | N/A | 3 | 3 | 3 | N/A |
| 2 | 4 | 1 | 5 | N/A | 3 | 2 | 5 | N/A |
| 3 | 3 | 3 | 8 | N/A | 1 | 1 | 6 | N/A |
| 4 | 2 | 0 | 8 | N/A | 1 | 1 | 6 | N/A |
| 5 | 3 | 2 | 10 | N/A | 2 | 0 | 6 | N/A |
| 6 | 1 | 0 | 10 | N/A | 2 | 1 | 7 | N/A |
| 7 | 1 | 1 | 11 | N/A | 2 | 1 | 8 | N/A |
| 8 | 1 | 0 | 11 | N/A | 1 | 0 | 8 | N/A |
| 9 | 1 | 0 | 11 | N/A | 0 | 0 | 8 | N/A |
| 10 | 1 | 0 | 11 | N/A | 1 | 0 | 8 | N/A |
| 11 | 3 | 0 | 11 | N/A | 1 | 0 | 8 | N/A |
| 12 | 1 | 0 | 11 | N/A | 1 | 2 | 10 | N/A |
| 13 | 1 | 0 | 11 | N/A | 1 | 1 | 11 | N/A |
| 14 | 1 | 0 | 11 | 0.0% | 1 | 0 | 11 | 10.0% |
| 15 | 1 | 0 | 11 | 0.0% | 2 | 0 | 11 | 0.0% |
| 16 | 1 | 0 | 11 | 0.0% | 0 | 0 | 11 | 0.0% |
| 17 | 1 | 0 | 11 | 0.0% | 2 | 2 | 13 | 20.0% |
| 18 | 1 | 0 | 11 | 0.0% | 1 | 0 | 13 | 20.0% |
| 19 | 1 | 0 | 11 | 0.0% | 1 | 0 | 13 | 0.0% |
| 20 | 2 | 0 | 11 | 0.0% | 2 | 1 | 14 | 10.0% |
| 21 | 1 | 0 | 11 | 0.0% | 1 | 0 | 14 | 10.0% |
| 22 | 1 | 0 | 11 | 0.0% | 4 | 2 | 16 | 20.0% |
| 23 | 2 | 1 | 12 | 9.1% | 2 | 1 | 17 | 30.0% |
| 24 | 1 | 0 | 12 | 9.1% | 3 | 2 | 19 | 30.0% |
| 25 | 2 | 0 | 12 | 0.0% | 3 | 1 | 20 | 30.0% |
| 26 | 0 | 0 | 12 | 0.0% | 1 | 0 | 20 | 10.0% |
| 27 | 1 | 1 | 13 | 9.1% | 2 | 1 | 21 | 10.0% |
| 28 | 2 | 1 | 14 | 18.2% | 1 | 1 | 22 | 20.0% |
| 29 | 2 | 0 | 14 | 9.1% | 2 | 0 | 22 | 10.0% |
| 30 | 1 | 0 | 14 | 0.0% | 1 | 0 | 22 | 0.0% |
| 31 | 2 | 0 | 14 | 0.0% | 2 | 0 | 22 | 0.0% |
| 32 | 2 | 0 | 14 | 0.0% | 2 | 0 | 22 | 0.0% |
| 33 | 2 | 1 | 15 | 9.1% | 1 | 0 | 22 | 0.0% |
| 34 | 0 | 0 | 15 | 9.1% | 1 | 1 | 23 | 10.0% |
| 35 | 0 | 0 | 15 | 0.0% | 1 | 0 | 23 | 10.0% |
| 36 | 0 | 0 | 15 | 0.0% | 1 | 0 | 23 | 0.0% |

Table 1 shows that two interviews in a row were completed with no new barriers identified by interview number 14. By this point, 11 barriers had been identified. In the remaining 22 interviews a further four barriers were identified.

For the solutions, saturation was achieved by interview number 15. By this point 11 unique solutions had been identified. By the end of 36 interviews, a further 12 solutions had been identified.

In summary, had we used Guest and colleagues approach with a conservative base of 12, less conservative runs of two, and a very conservative new information threshold of 0%, we would have stopped after interview number 15, missing out on a third of the unique barriers that emerged across our 36 telephone interviews.

In our next analysis, we counted the total number of barriers *and* solutions (themes) that emerged from each interview. We ran six additional scenarios, using runs of two, three, and four against the base of all themes identified in the first 12 interviews. We also used runs of two, three, and four against all themes identified in all preceding interviews.

Table 2 shows that using a more conservative base that includes all interview conducted does not materially change the point at which saturation is deemed to have been achieved. However, increasing the run length from three to four was a associated with a much later saturation point; 32 interviews with a 0% new information threshold. Interestingly, the use of a higher new information threshold (<5%) would lead to very similar saturation points of between 14-16 interviews depending on run length.

**Table 2:** Different run lengths and base sizes to assess saturation, using all themes in each interview.

| Interview number | Themes identified | Novel themes | Cumulative themes | Runs of 2 |  | Runs of 3 |  | Runs of 4 |  |
| --- | --- | --- | --- | --- | --- | --- | --- | --- | --- |
|  |  |  |  | Base = 12 | Base ≥12 | Base = 12 | Base ≥12 | Base = 12 | Base ≥12 |
| 1 | 7 | 7 | 7 |  |  |  |  |  |  |
| 2 | 7 | 3 | 10 |  |  |  |  |  |  |
| 3 | 4 | 4 | 14 |  |  |  |  |  |  |
| 4 | 3 | 1 | 15 |  |  |  |  |  |  |
| 5 | 5 | 2 | 17 |  |  |  |  |  |  |
| 6 | 3 | 1 | 18 |  |  |  |  |  |  |
| 7 | 3 | 2 | 20 |  |  |  |  |  |  |
| 8 | 2 | 0 | 20 |  |  |  |  |  |  |
| 9 | 1 | 0 | 20 |  |  |  |  |  |  |
| 10 | 2 | 0 | 20 |  |  |  |  |  |  |
| 11 | 4 | 0 | 20 |  |  |  |  |  |  |
| 12 | 2 | 2 | 22 |  |  |  |  |  |  |
| 13 | 2 | 1 | 23 |  |  |  |  |  |  |
| 14 | 2 | 0 | 23 | 4.5% | 4.5% |  |  |  |  |
| 15 | 3 | 0 | 23 | 0.0% | 0.0% | 4.5% | 4.5% |  |  |
| 16 | 1 | 0 | 23 | 0.0% | 0.0% | 0.0% | 0.0% | 4.5% | 4.5% |
| 17 | 3 | 2 | 25 | 9.1% | 8.7% | 9.1% | 8.7% | 9.1% | 8.7% |
| 18 | 2 | 0 | 25 | 9.1% | 8.7% | 9.1% | 8.7% | 9.1% | 8.7% |
| 19 | 2 | 0 | 25 | 0.0% | 0.0% | 9.1% | 8.7% | 9.1% | 8.7% |
| 20 | 4 | 1 | 26 | 4.5% | 4.0% | 4.5% | 4.0% | 13.6% | 13.0% |
| 21 | 2 | 0 | 26 | 4.5% | 4.0% | 4.5% | 4.0% | 4.5% | 4.0% |
| 22 | 5 | 2 | 28 | 9.1% | 7.7% | 13.6% | 12.0% | 13.6% | 12.0% |
| 23 | 4 | 2 | 30 | 18.2% | 15.4% | 18.2% | 15.4% | 22.7% | 20.0% |
| 24 | 4 | 2 | 32 | 18.2% | 14.3% | 27.3% | 23.1% | 27.3% | 23.1% |
| 25 | 5 | 1 | 33 | 13.6% | 10.0% | 22.7% | 17.9% | 31.8% | 26.9% |
| 26 | 1 | 0 | 33 | 4.5% | 3.1% | 13.6% | 10.0% | 22.7% | 17.9% |
| 27 | 3 | 2 | 35 | 9.1% | 6.1% | 13.6% | 9.4% | 22.7% | 16.7% |
| 28 | 3 | 2 | 37 | 18.2% | 12.1% | 18.2% | 12.1% | 22.7% | 15.6% |
| 29 | 4 | 0 | 37 | 9.1% | 5.7% | 18.2% | 12.1% | 18.2% | 12.1% |
| 30 | 2 | 0 | 37 | 0.0% | 0.0% | 9.1% | 5.7% | 18.2% | 12.1% |
| 31 | 4 | 0 | 37 | 0.0% | 0.0% | 0.0% | 0.0% | 9.1% | 5.7% |
| 32 | 4 | 0 | 37 | 0.0% | 0.0% | 0.0% | 0.0% | 0.0% | 0.0% |
| 33 | 3 | 1 | 38 | 4.5% | 2.7% | 4.5% | 2.7% | 4.5% | 2.7% |
| 34 | 1 | 1 | 39 | 9.1% | 5.4% | 9.1% | 5.4% | 9.1% | 5.4% |
| 35 | 1 | 0 | 39 | 4.5% | 2.6% | 9.1% | 5.4% | 9.1% | 5.4% |
| 36 | 1 | 0 | 39 | 0.0% | 0.0% | 4.5% | 2.6% | 9.1% | 5.4% |

Finally, we plotted the cumulative themes that emerged over the course of the 36 interviews. Figure 1 illustrates the fact that the majority of unique barriers were identified by interview number seven, followed by a long plateau. Shorter plateaus in the number of solutions arising from interviews 7-11 and 13-16 were followed by incremental gains until levelling off again around interview number 28. The paucity of new information arising from interviews 13-16 drive the saturation decisions in the analyses performed above.

**Figure 1:**
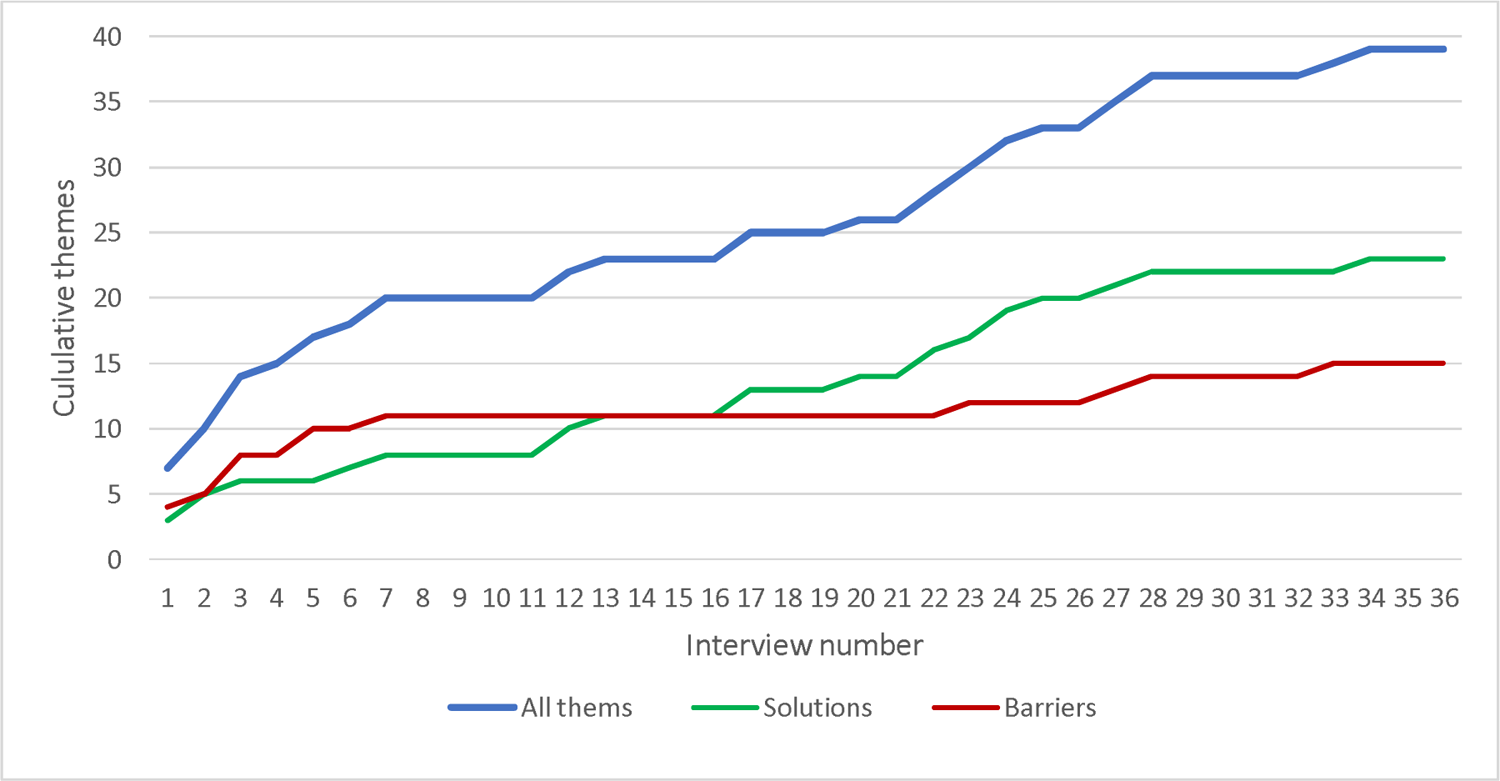
Accumulation of themes as the interviews progressed

Figure 1 suggests that for this specific set of interviews, a base size of 15 and a run length of three might have yielded a more appropriate saturation point, given that new themes continued to accrue until around interview number 28. Table 3 shows that using a conservative base of ≥15 interviews and a new information threshold of 0% leads to saturation by interview number 30, 31, and 32 with respective run lengths of two, three, and four.

**Table 3:** Re-running the saturation analysis with a base of => 15 and runs of 2-4.

| Interview number | New information with a base of $\geq 15$ | | |
| --- | --- | --- | --- |
|  | *Runs of 2* | *Runs of 3* | *Runs of 4* |
| 15 |  |  |  |
| 16 |  |  |  |
| 17 | 8.7% |  |  |
| 18 | 8.7% | 8.7% |  |
| 19 | 0.0% | 8.7% | 8.7% |
| 20 | 4.0% | 4.0% | 13.0% |
| 21 | 4.0% | 4.0% | 4.0% |
| 22 | 7.7% | 12.0% | 12.0% |
| 23 | 15.4% | 15.4% | 20.0% |
| 24 | 14.3% | 23.1% | 23.1% |
| 25 | 10.0% | 17.9% | 26.9% |
| 26 | 3.1% | 10.0% | 17.9% |
| 27 | 6.1% | 9.4% | 16.7% |
| 28 | 12.1% | 12.1% | 15.6% |
| 29 | 5.7% | 12.1% | 12.1% |
| 30 | 0.0% | 5.7% | 12.1% |
| 31 | 0.0% | 0.0% | 5.7% |
| 32 | 0.0% | 0.0% | 0.0% |
| 33 | 2.7% | 2.7% | 2.7% |
| 34 | 5.4% | 5.4% | 5.4% |
| 35 | 2.6% | 5.4% | 5.4% |
| 36 | 0.0% | 2.6% | 5.4% |

Researchers like Coenen et al. have suggested that run lengths of two are adequate,[3] whilst others have recommended runs of three.[2,4] Based on our findings presented in table we decided that we would use a base of ≥15 and runs of three for future studies.

### Application to second data set

As we had conducted a second set of 31 in-person interviews, we were able to test whether the ≥15+3 approach would have ended data collection at an appropriate point, i.e. without missing additional themes that arose in further interviews, nor prolonging data collection long after all themes had been identified.

Table 4 shows that using this approach identifies interview number 24 as the point at which saturation is achieved.

**Table 4:**
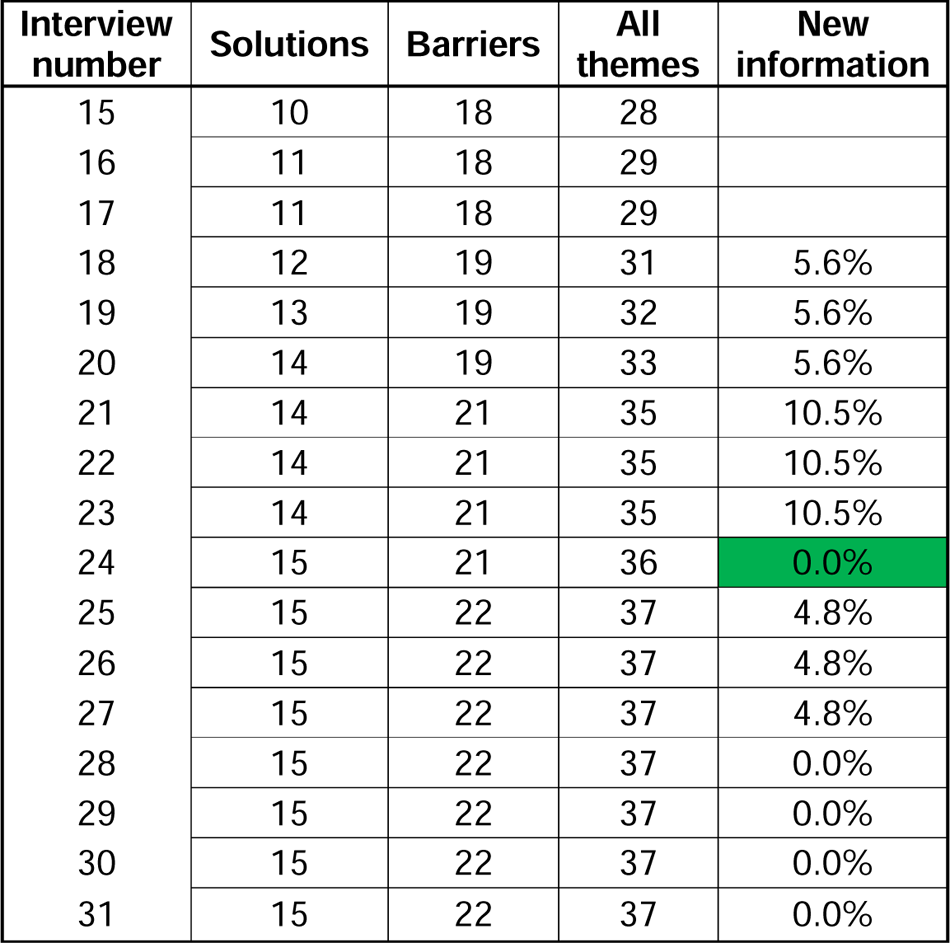
Saturation analysis of 31 in-person interviews using a base of ≥15 and a run length of three.

Using this cutoff would have led to the inclusion of all solutions, and all but one of the barriers identified across the 31 interviews. Figure 2 plots the saturation point on a line graph.

**Figure 2:**
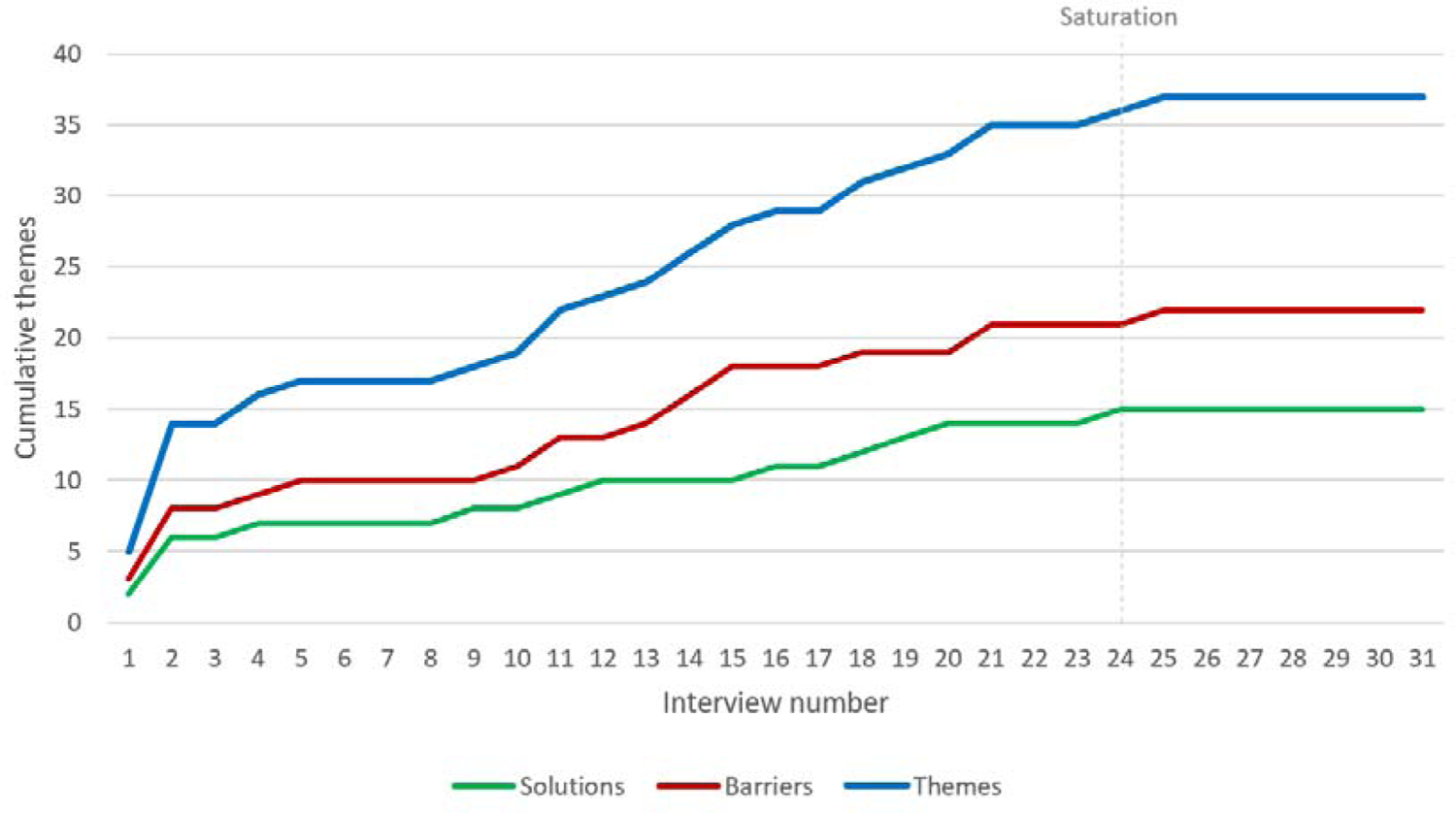
Accumulation of themes as the interviews progressed

Based on this retrospective analysis of our data, our group has decided to use the 15+3 approach to assess saturation in our future qualitative research projects.

## Appendix 5

**Supplementary Table 1:**
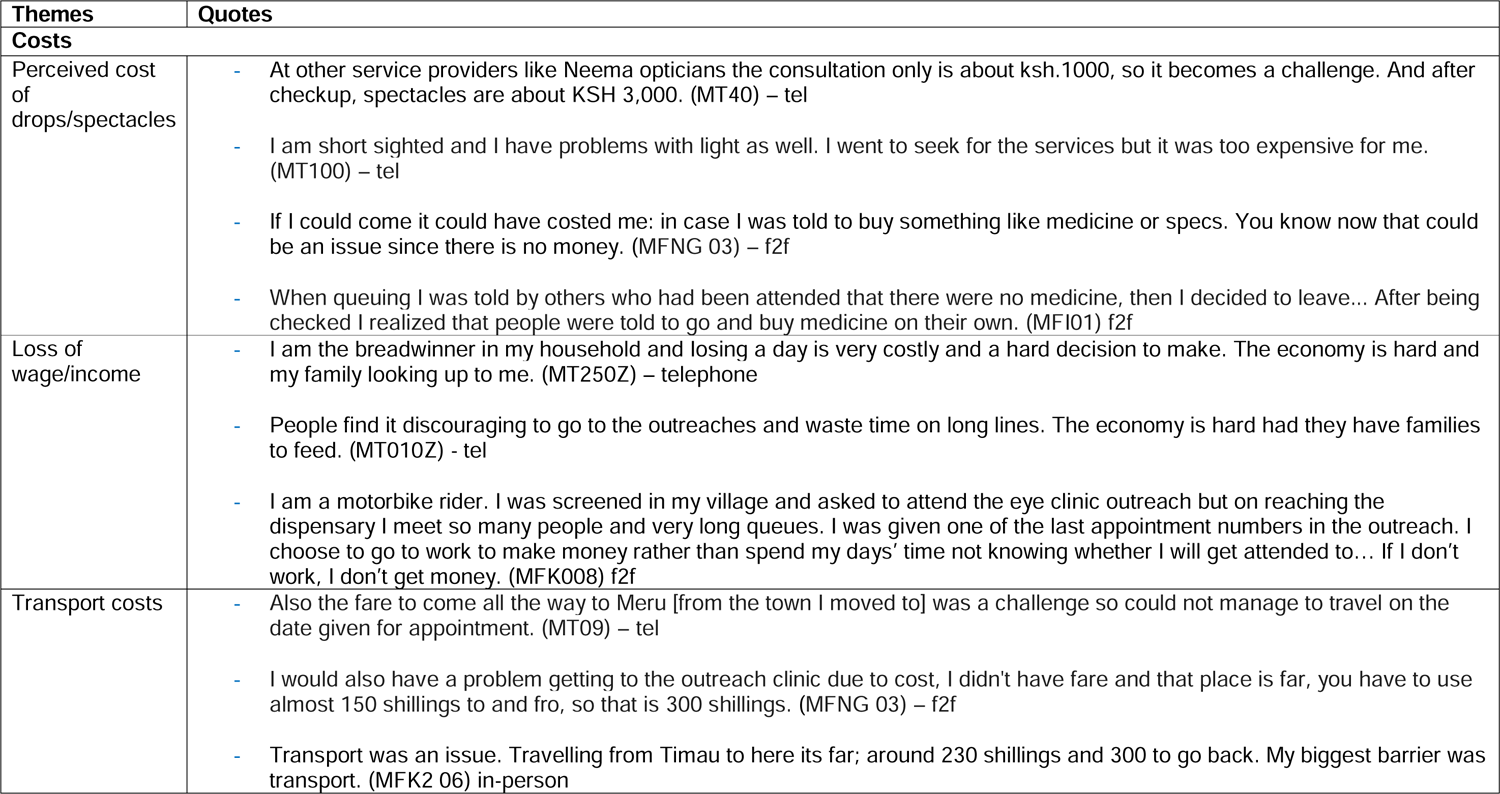
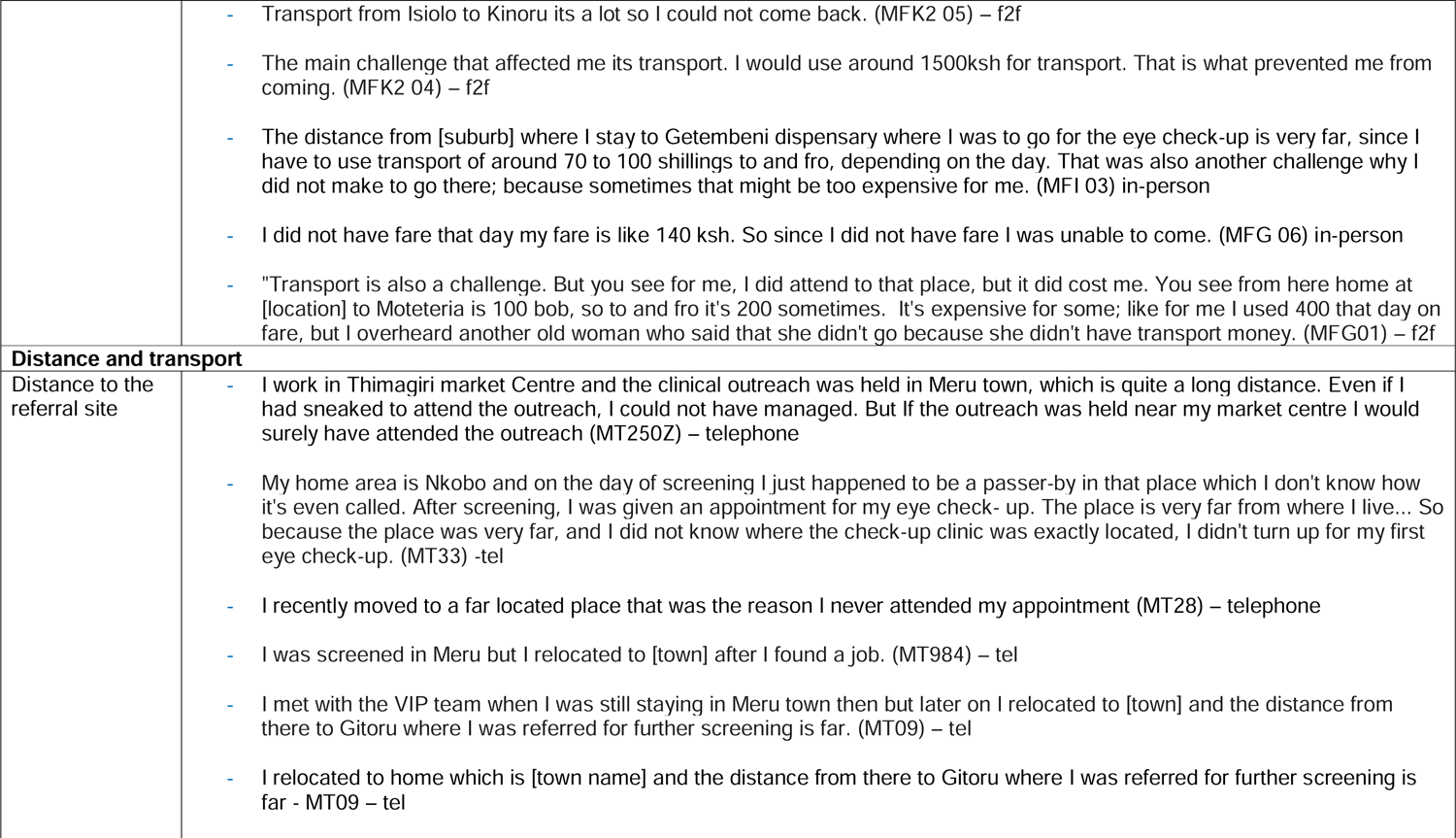
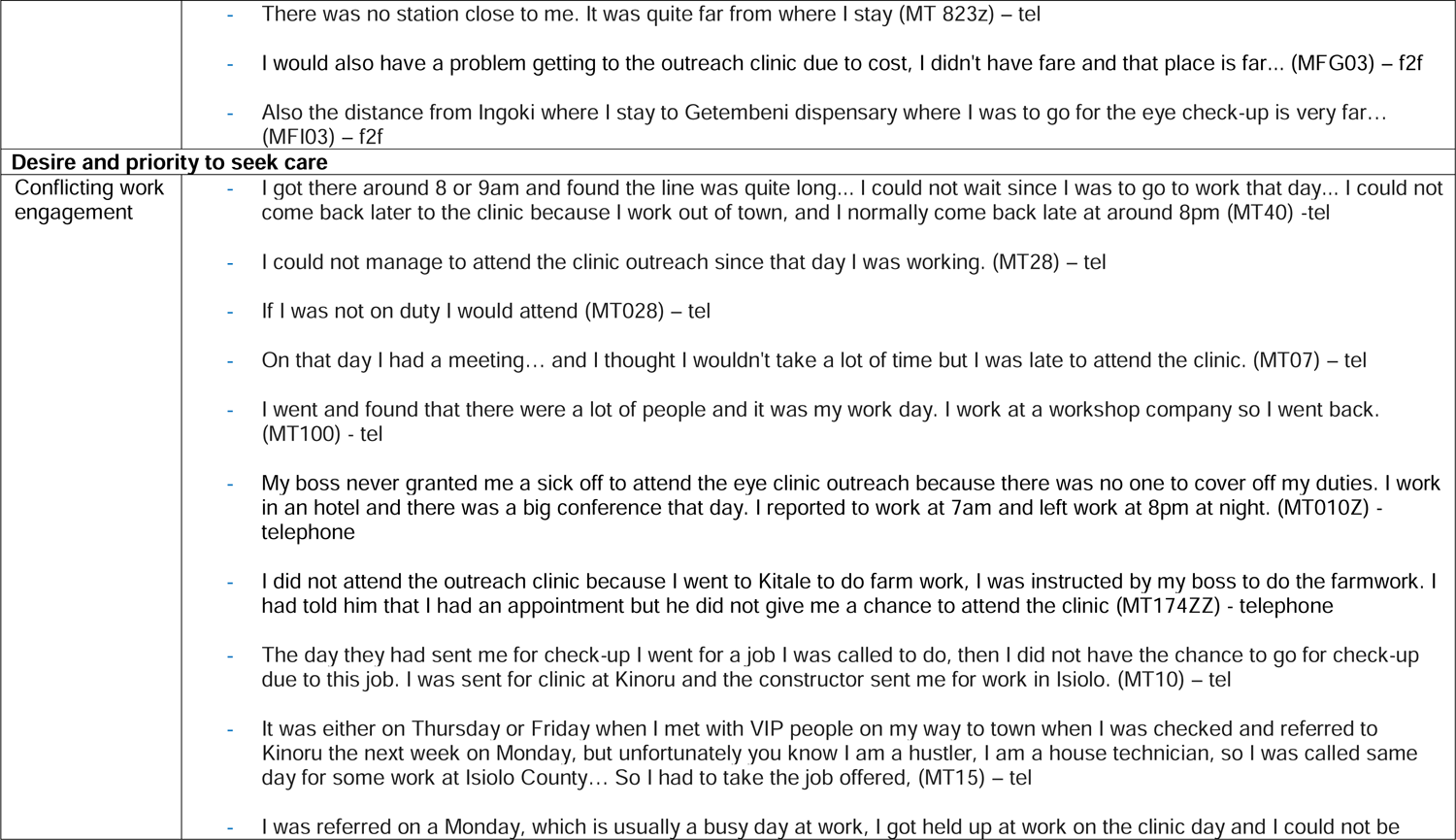
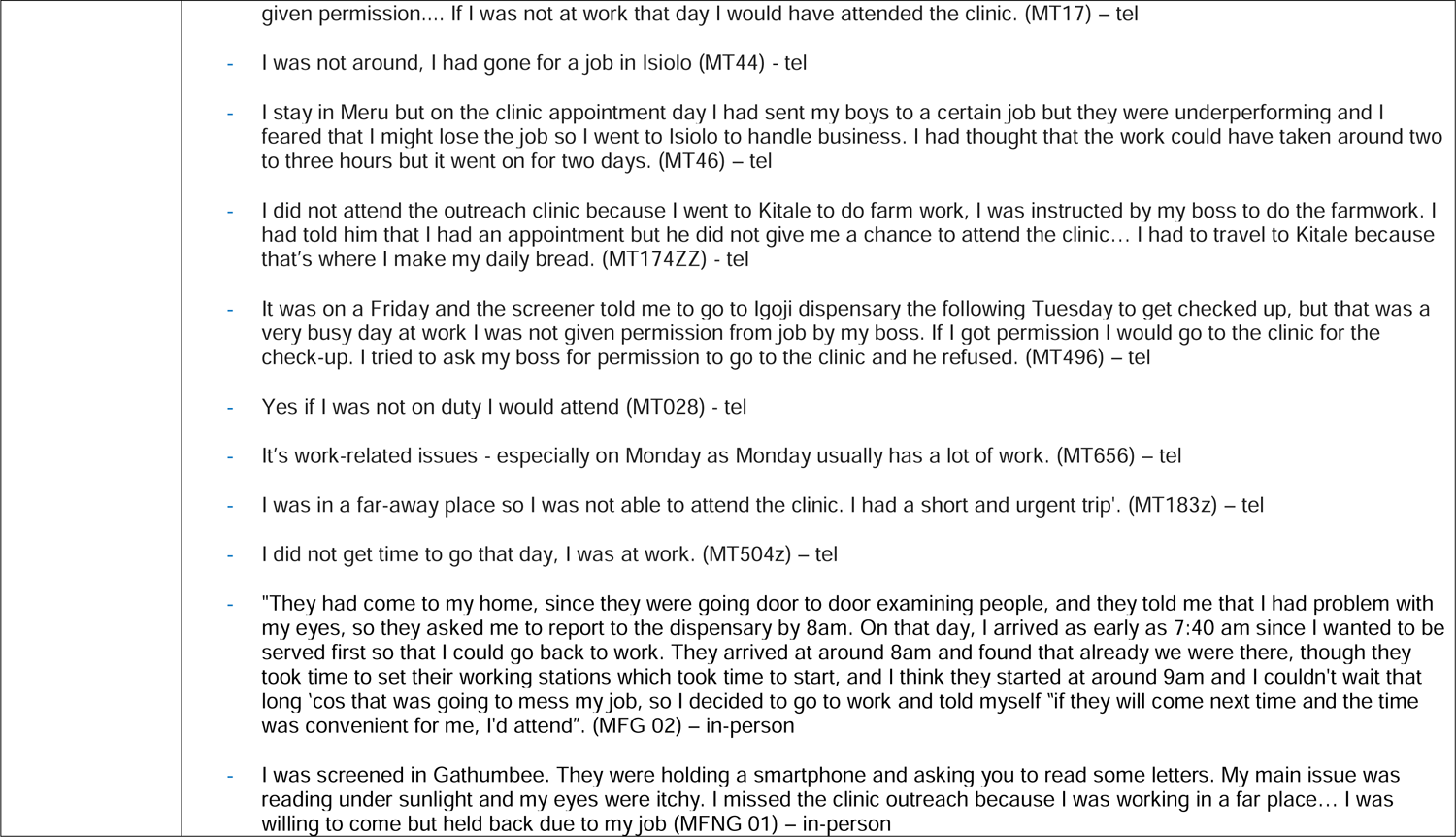
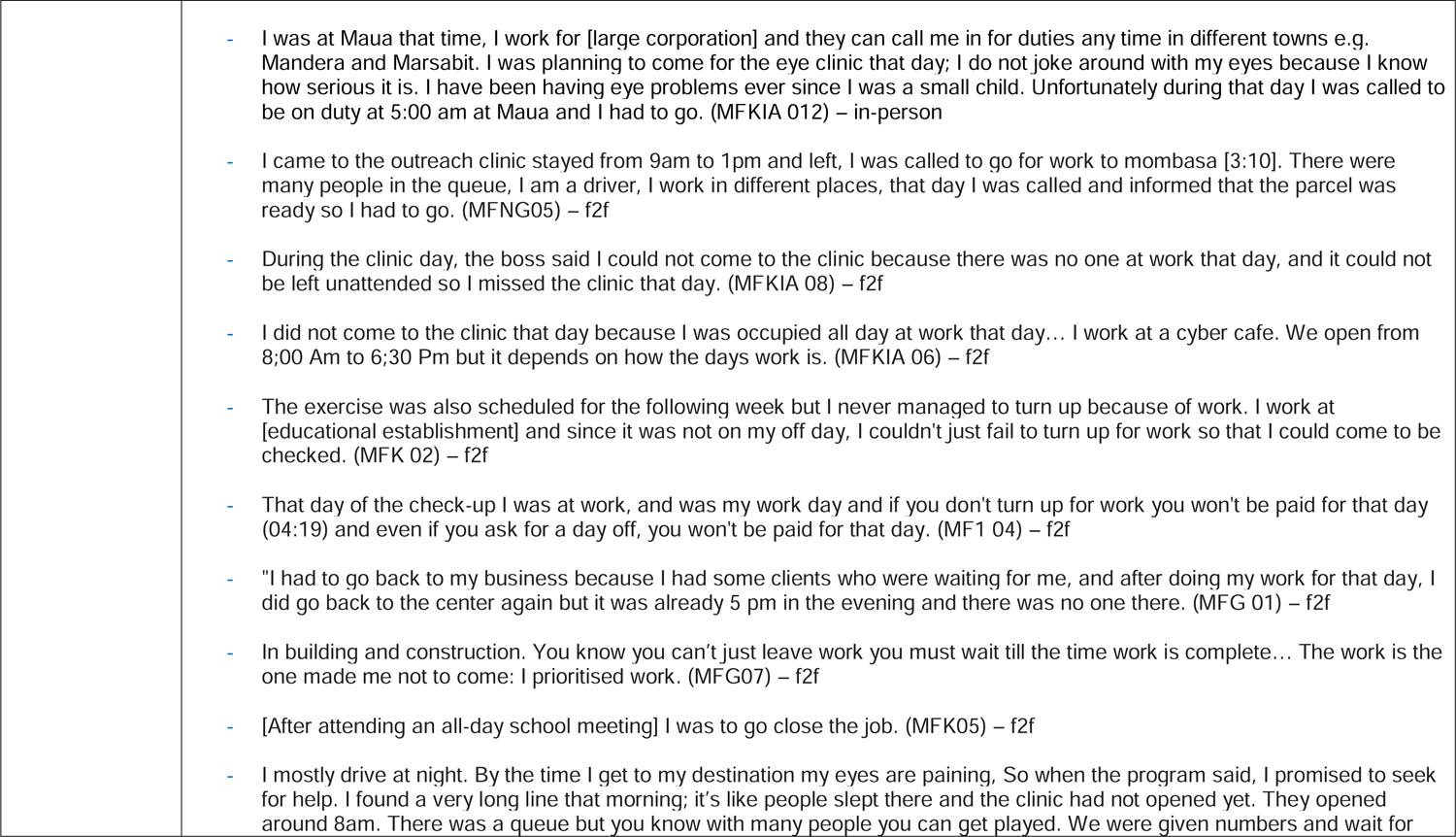
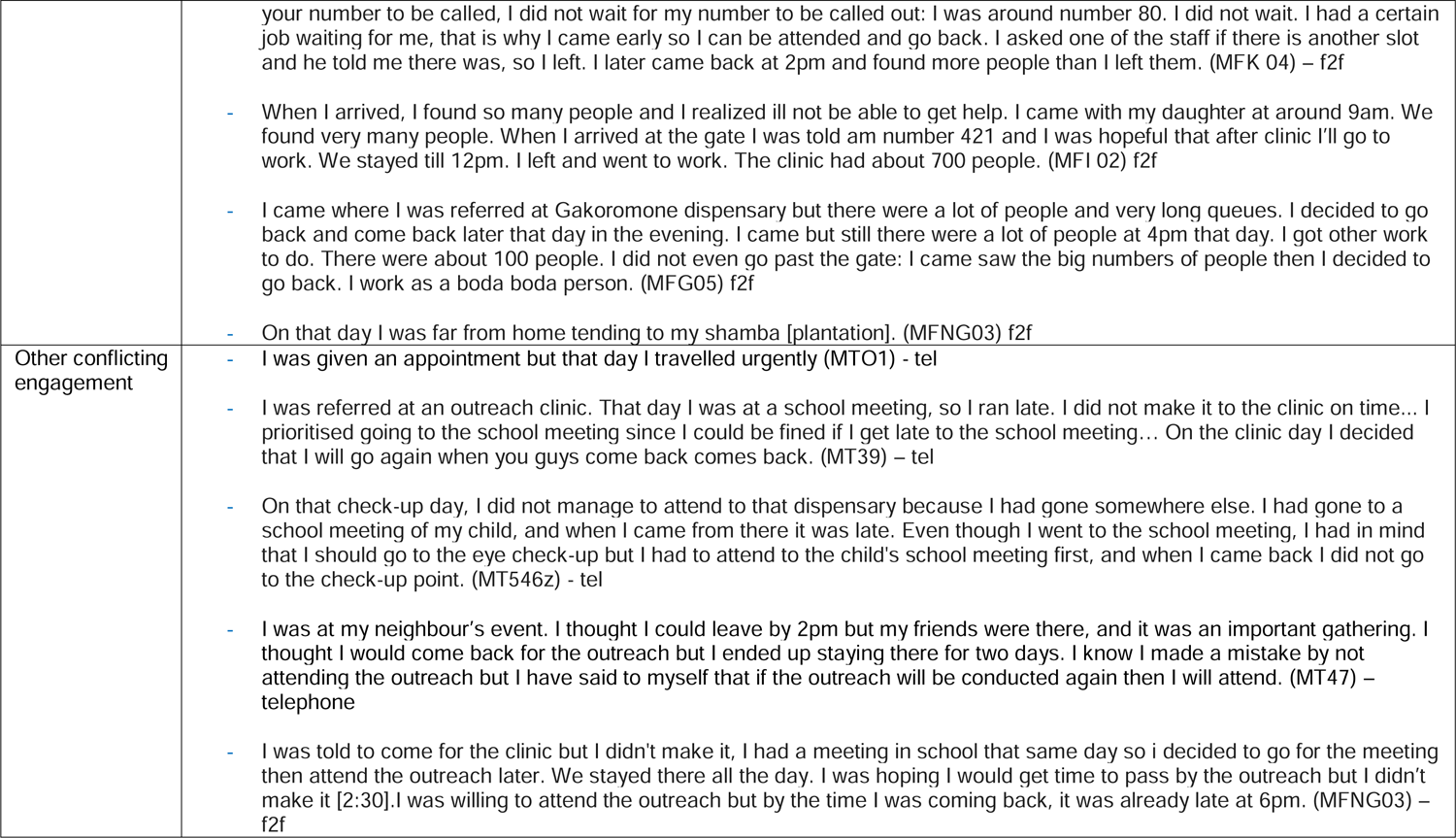
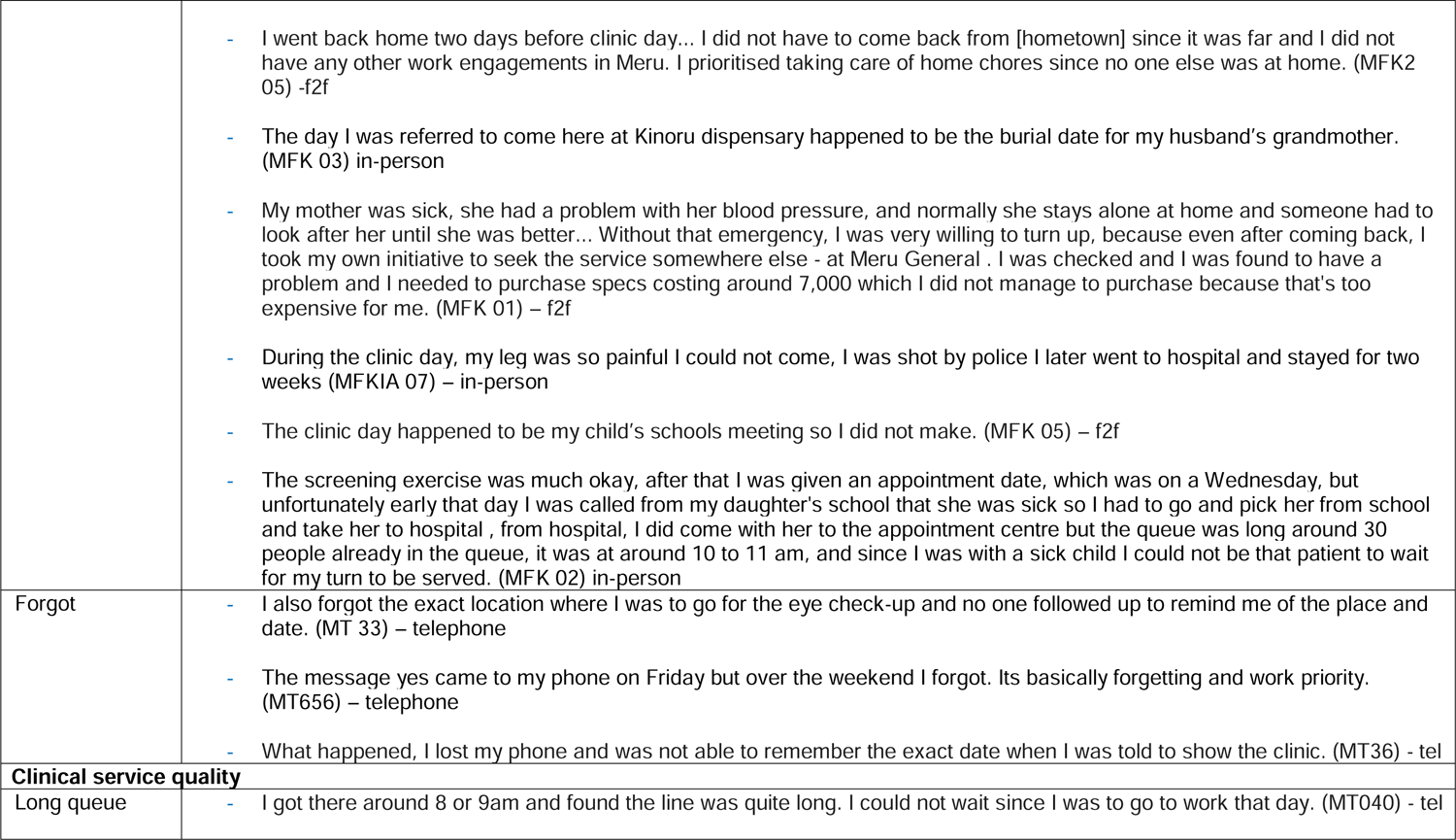
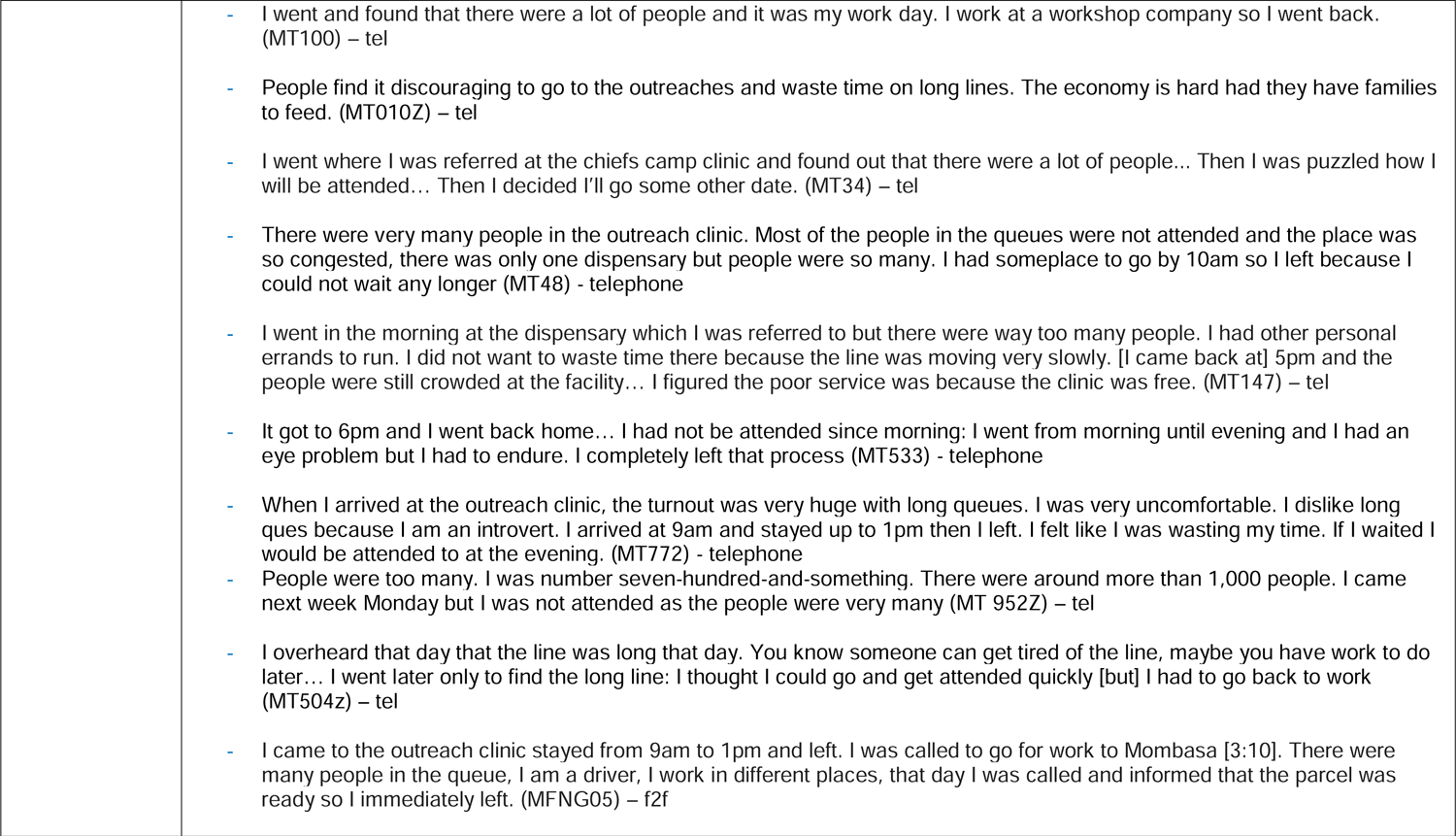
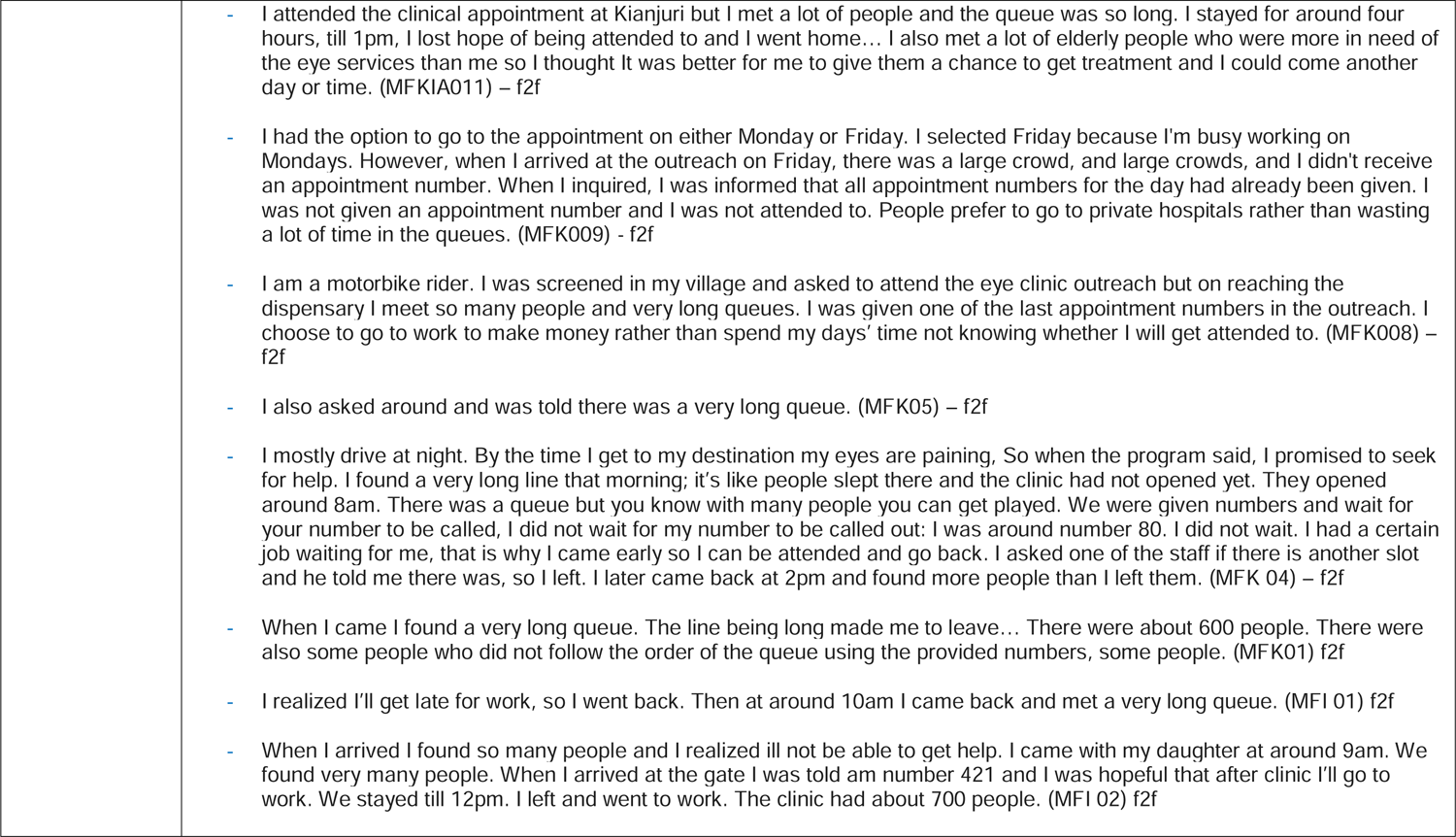
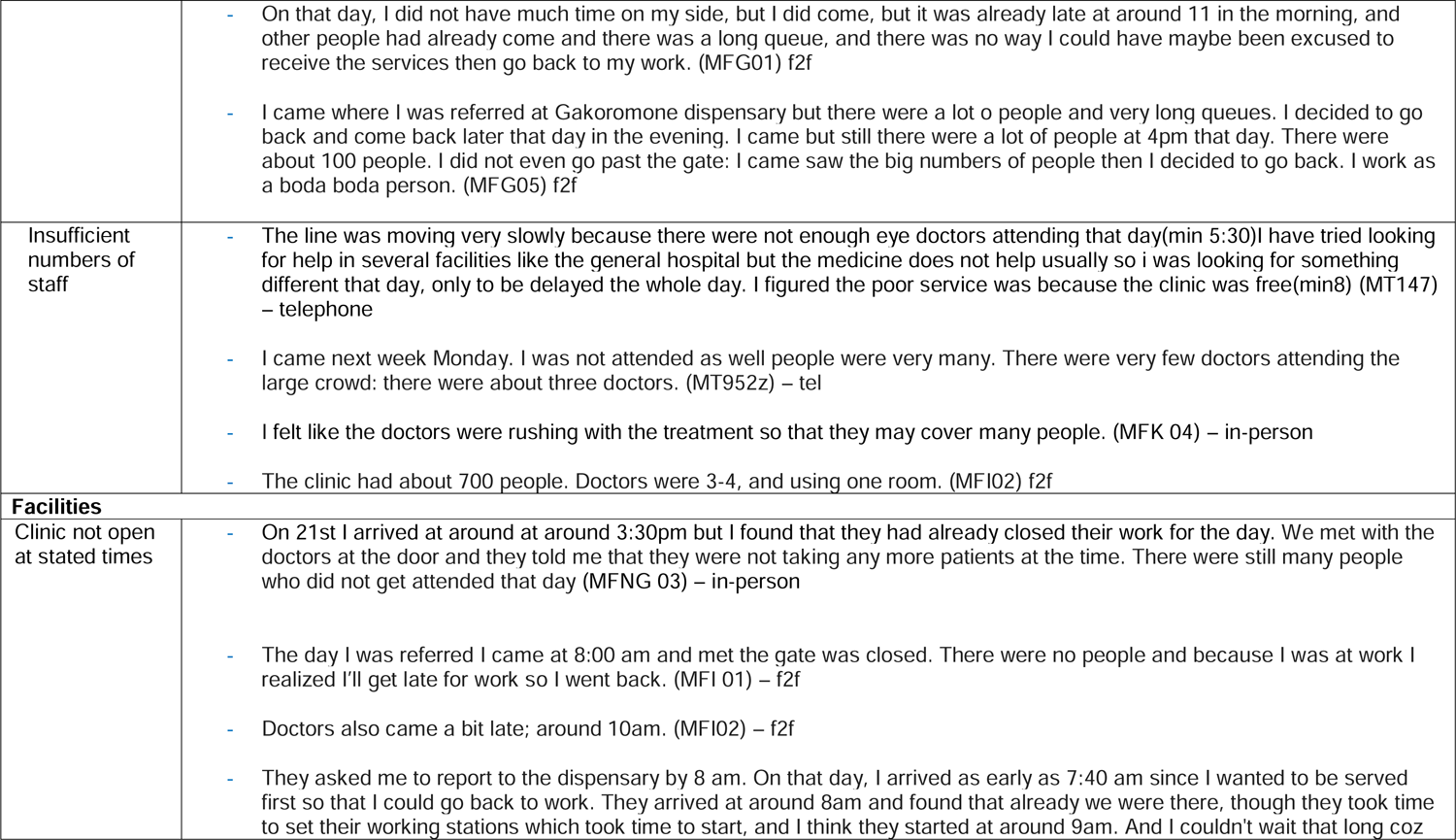
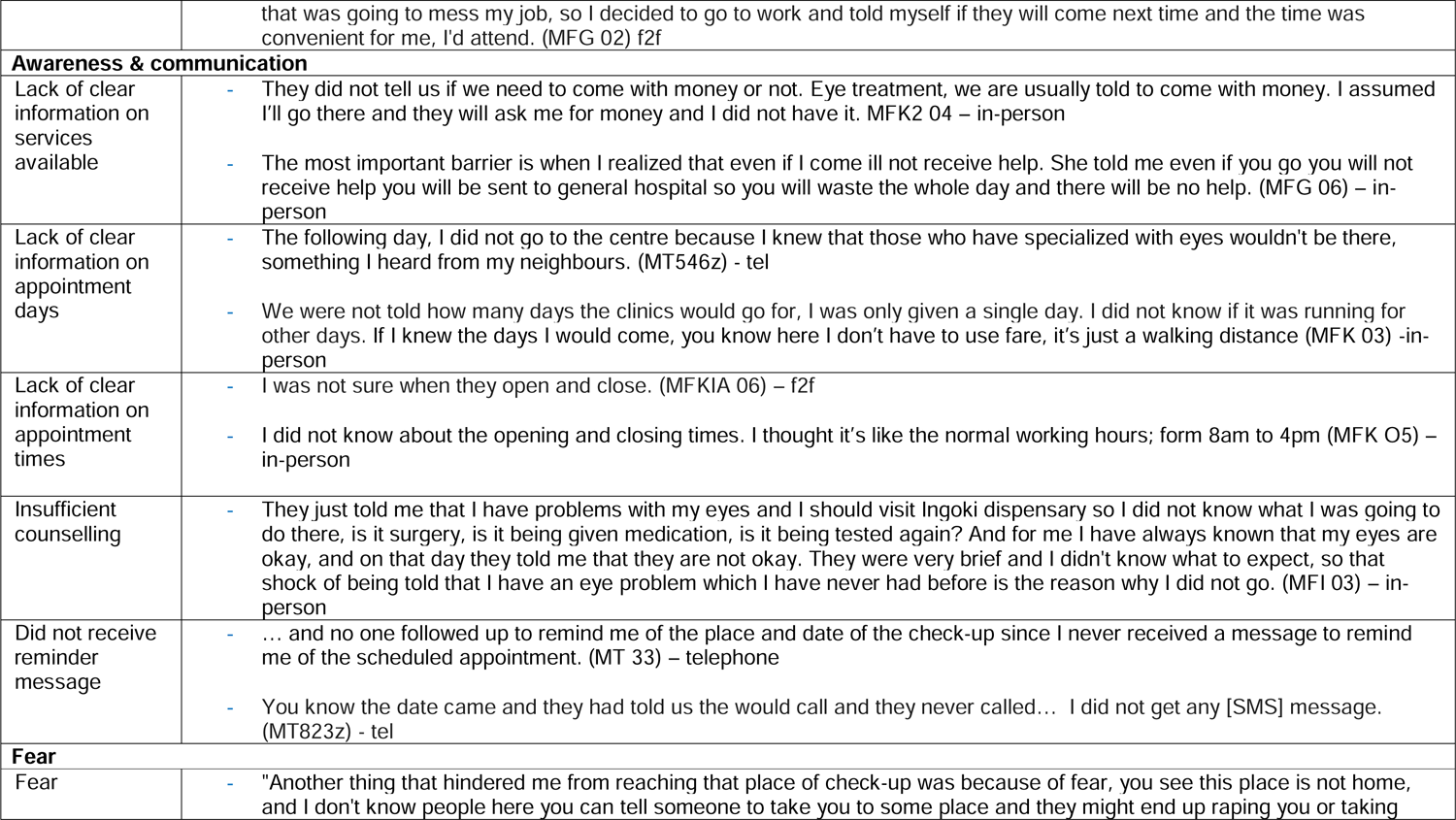
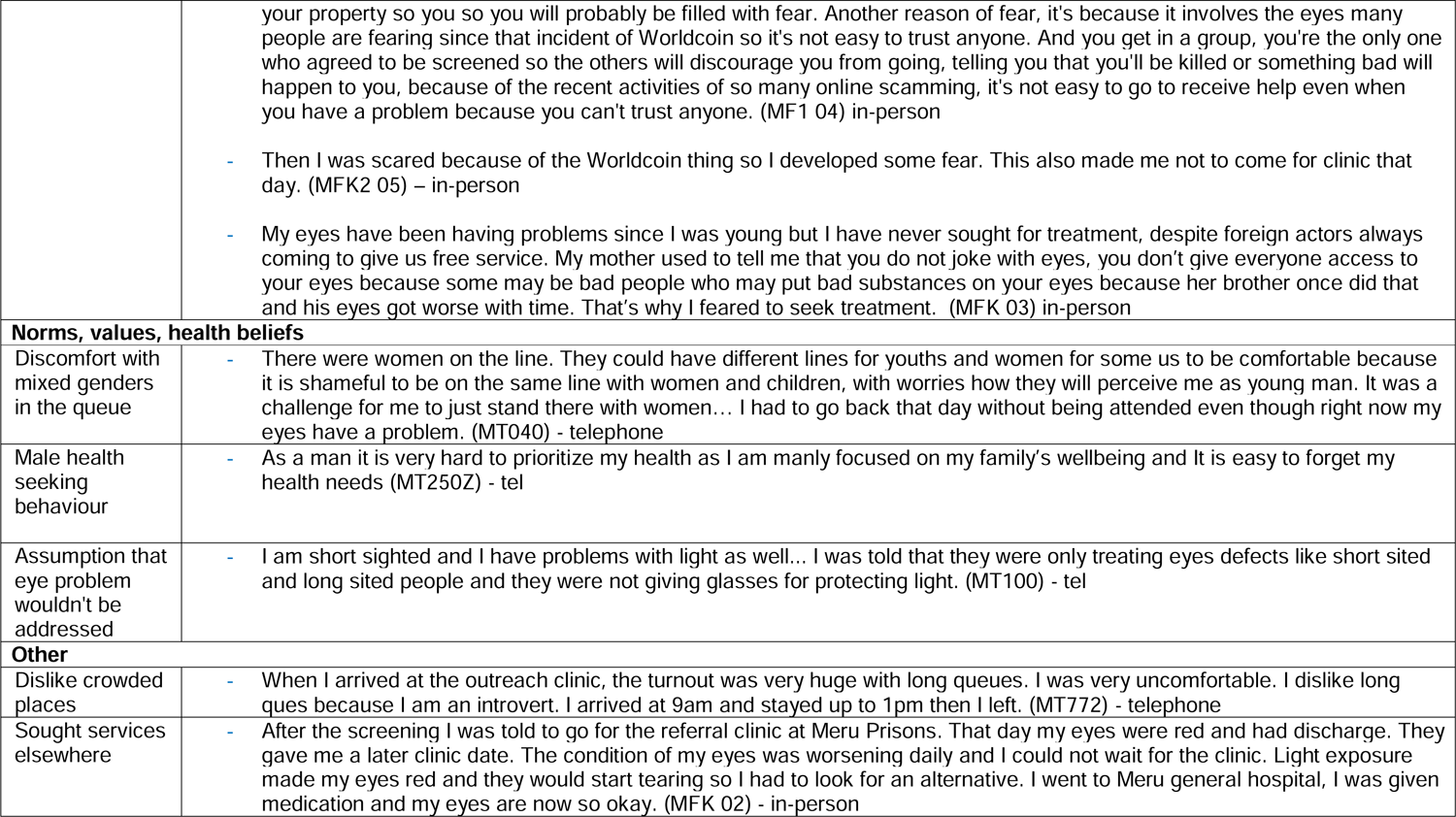
Barrier quotes.

## Appendix 6

**Supplementary Table 2:** Solution quotes.

Appendix 6: Supplementary Table 2: Solution quotes
| Theme | Quotes |
| --- | --- |
| **BARRIERS** |  |
| **Costs** |  |
| Provide transport fare | If I can be provided with fare, I would definitely attend the clinic because the challenge is income and moving to Gitoru requires money - MT 09 – telephone<br>According to how the economy is, if we can be given fare, at least you can borrow some money for transport to attend the outreach, then refund later. You can also help people after screening at least give some assistance, you can tell us to look for some money and then you top up so that we can manage to buy medicine and specs since if its only coming to be checked, even if I keep coming again and again it will not be of much help. MFNG 03 –<br>I also feel like, at least we should be provided with fare - MFG 01 – |
| Subsidise treatment | You should consider needy people who cannot afford [spectacles]... At least organise yourselves and look for the needy to help them, If I get that money I can go and get the spectacles because no one is happy when their bodies have a problem. I cannot even read the bible or the numbers on the phone - MT 148 – tel<br>According to how the economy is, if we can be given fare, at least you can borrow some money for transport to attend the outreach, then refund later. You can also help people after screening at least give some assistance, you can tell us to look for some money and then you top up so that we can manage to buy medicine and specs since if its only coming |
|  | to be checked, even if I keep coming again and again it will not be of much help. MFNG 03 – |
| Pay people to attend | You should also give people some little money if they attend the appointment - MT 46 - tel<br>I'm asking if we can be compensated with the amount we could have been paid because we leave our job so at least you are sure - even if you're not going to work on that day - your children are not going to sleep hungry. Because some of us, we're the providers of our families. MFI 04 – |
| ***Distance and transport*** |  |
| Move clinics closer to where people live and work | The outreach should be situated in a place which is easily accessible to many people and where many people live... It should not be very far from where people live - MT 28 – tel<br>Because I was just a passerby, it will be better if the eye screening was brought to where I live in Nkobo – MT33 – tel<br>Have the stations close to the people where we can all reach. MT 823z – tel<br>I can go to the outreach clinic if its situated near me where i will get help but not going all the way to Meru MT682 - tel<br>If the outreach was held near my market centre I would surely have attended the outreach – MT 250Z – tel<br>You should bring the services closer to the people... Because you have the people's contacts and where they live, you can conduct an outreach in a place which near to where they live. MT 183z - tel<br>They should bring the clinics near the people to cut cost on transport - MFK2 04 – |
|  | Services should also be near - some facilities are at a long distance so it becomes difficult to go there. - MFK 009 – in-person<br>For distance, I think you should organize and bring the services to someplace that's near to us for example chief's camp - MFI 03 – in-person<br>Clinics should be set in various places in every county. So that the clinics would be near people. MFK2 05 – in-person<br>I am suggesting if they could take the services to every dispensary and for my case, a near dispensary like for me, [name] dispensary. - MFG 01- in-person<br>Also, it would be easier if they could bring the services to nearer facilities for instance for people from my area, to [name] dispensary - MFG 02 – in-person |
| Provide transport | If you give me transport and day to come, I will come. MT 10 – telephone<br>For the elderly you can look for means of transport, many of them will come. You can organise with people like chiefs so that they can know how to take people to the places where the services are offered. - MFK 009 – in-person<br>if they would be able to provide transport that would be OK. MFG 07 – in-person |
| Door-to-door services | Is it possible to have a doctor visiting them [elderly people] door to door? It would be better for those who aren't able to go there - MT 15 – tel |
|  | If possible, you can take the services to people's homes considering there are those very old who cannot make it to the appointment centres - MFK 01 – in-person<br>Consider door to door services but I don't know if that's possible. MFI 03 – |
| **Clinical service quality** |  |
| Improve punctuality and speed of staff | I think those that are providing the services should be punctual and fast - MT 07 – tel<br>Increase [the number of] doctors from four to ten, and then those doctors to arrive early at around 8am– MFI 02 - in-person<br>The solution is still time, though 8am is still okay, but I think if maybe they could work on their organization so that much time is not wasted on preparing the working stations - MFG 02 – in-person |
| Ensure that all medicines and glasses can be provided on-site | Equip more drugs in the dispensary - MT 48 – tel<br>When you see someone, make sure they get medication or give them spectacles; not writing prescriptions and referring them to chemists. I think this does not help. You should have all the medication for all allergies and spectacles, not referring people to places - MT 533 – tel<br>After being screened you should receive all the services like medicine and everything, you get solution for everything but not again looking for services from other places. MFNG05 – in-person |
|  | Also, the program should improve on their equipment and machines used for examination, so that the services will be satisfactory. MFG 01 – in-person<br>Next time let the doctors be many and let them come with medicines. MFI01 – in-person<br>For the issue of specs , maybe you can add more resources that are enough for everyone next time. MFG03 – in-person |
| Add more staff at each clinic | Add more eye doctors that day so that they may help each other in covering more people on the clinic days - MT 147 – tel<br>Have many check-up doctors, not bringing two or three of them. That day they were only five or six, you see them attending all those people - is impossible - MT 533 – tel<br>Increase the number of doctors so that the clients can be served more efficiently and on time - MT 772 – tel<br>Add more doctors so that everybody will be served - MT 952Z - tel<br>Maybe you should add more doctors to attend to more people so the lines can move faster - MT 504 Z – tel<br>Those people doing screening should be many because staying for two three hours with this economy is a loss. MFNG05 – in-person |
|  | There is a high number of people requiring eye services thus the number of doctors should increase - MFKIA 011 – in-person<br>Increase the staff because there were very many people increase the doctors from four to ten - MFK2 04 – in-person<br>Increase doctors from four to ten. MFI 02 – in-person<br>Increase no of doctors so that I will not take long waiting because I have other duties to do. Next time let the doctors be many and let them come with medicine... Add doctors from three to six: that would be OK. – MFI 01 – in-person<br>The doctors attending people should be many. Because like... people were so many, and most of them went back without being attended to. So if there were many doctors it would have been easy. MFK 009 – in-person<br>I think next time you should increase the number of doctors attending so that every individual may get good quality service - MFK 05 – in-person<br>Maybe next time you can add more manpower because for the long queues and stuff... I have a feeling that there are those who never got attended to at all because of lack of enough time, due to inadequate staff - MFG 03 – in-person<br>...and set more stations, and also doctors to be added in those stations. MFG 05 – in-person |
| Schedule fewer people to attend each day | When you register, register few people... Having 1000 people wastes a lot of time and you cannot handle such a crowd. If you target 100 people, you will attend all of them. Having many people will have them trolling around all day. You see, |
|  | I had an eye problem but I swore never to come back again, I would rather go to the hospital than come waste time around just because it is free - MT 533 – tel<br>The outreach should limit the number of clients they are attending to. It make no sense to refer 600 people and they cannot fully attend to in a day. They should choose a much smaller number comprised of people they had referred. MT 772 – tel<br>You should take time to examine every patient as it should without minding the workload ahead, that's why I think you should schedule less people. MFK04 – in-person<br>I would also like it if you control the number of people you schedule for a particular day, because now you have experience of how many people you can attend for a day. Schedule just that amount of people so that everyone may be attended to... It wasn't fair of you to call that many people, they were already sick and they had to wait for hours under the scorching sun. MFK01 – in-person |
| **Facilities** |  |
| Keep the clinics open for a greater number of days | The screening exercise should not be done just once, it should be repeated maybe even for three days during weekdays before an appointment for the first check-up is made - MT 33 – tel<br>Maybe you can add more days so that you can schedule few people on one day - MT 147 – tel<br>You should extended the number outreach days to attend to more people - MT 001 – tel |
|  | If there were more days I could have attended the clinic. I suggest that you add more clinic days - MT 39 - tel<br>Come at least two days consecutively; let it not be one day. If I miss one day, I will be able to go the following day - MT 34 – tel<br>Add more days for the clinic so that people who did not manage to attend the clinic on their appointment dates can still get a chance to be treated. Many people have eye problems but they are also busy looking for ways of surviving during these tough economic times - MT 174Z – tel<br>You should extended the number outreach days to attend to more people - MT 01 – tel<br>Also you should add the number of outreach days to attend to more people. MT47 - tel<br>Add more clinic days around 3-4 days - MT 952Z – tel<br>For me I'd suggest that next time you have a similar exercise, at least add more days because this time you had only specified for one day, this will assist those who hadn't managed to reach that day to have another chance. MT 546z - tel<br>Add more days. You see I was referred for one day if there was a next day I would purpose to come - MT 504 Z – tel<br>Having a single day for clinics gets people congested... I think that Monday, Wednesday, or even the whole week of clinic will be easier for most people to access the clinic as it is easier to ask for permission from work and be sure you |
|  | will be attended. With the whole week one can get a day to go and if they do not get attended that day, they can still choose the next day and will be sure to be attended. MT040 – tel<br>You should also do those outreach clinics for more days at least two or three days in a month. That time they were doing the outreach for just one day. MFNG05 – in-person<br>The outreach should not be in one day. It should be at least two days in a week. If you allow everyone to the outreach there will be long queues and many of them will not be attended. MFKIA 012 – in-person<br>You can also add days, if I miss today, I can come the next day. If you add the days, and make sure you remind us, it is okay. MFKIA 08 – in-person<br>If you add the clinic days I feel it is okay. MFKIA 07 – in-person<br>You can just extend the clinic days to have more of them, like four to five days. MFKIA 06 – in-person<br>More appointment days maybe for like 2 weeks. MFK 01 – in-person<br>If possible you can also add some days like then you were checking people on Monday and Friday because many people were interested but did not get help. MFK 009 – in-person<br>If it is possible, have the clinics for at least two days: if you miss today, you can purpose to come the following day |
|  | because even those people were too many. MFK 05 – in-person<br>You know if you give the clinic like three days, you can even travel for a safari, come back the next day and still attend the clinic. So I would suggest you schedule more days for the clinics. MFK 03 – in-person<br>Look for a day like Friday because work load is few. MFK04 – in-person |
| Extend clinic opening hours | If they would extend services to the evening at 5pm that would be okey since most people are out of work. MT100 – tel<br>I would say that you guys had a great job in our community according to many testimonies from the people who made it to the clinic and I regret not going, I feel devastated. However I would suggest that you add some more opening and closing hours so that people who are committed during the day can access the clinic. MT39 – tel<br>Open the clinics early and have the two days and people coming early. MFK 05 – in-person<br>Also have the clinics start as early, like if we come around 6:30am the clinics can open at 7am. I feel like that days clinic started late they started at 8am, I was there at 6:30am. MFK04 – in-person<br>Also if it's on weekdays, if it's possible if the exercise could extend the working hours, let's say from 4 pm to 6 pm, that one too I could manage, because I would have come from work. MFK02 – in-person |
| Keep the clinics open on the weekend | The biggest challenge for me was timing. If timing was ok I would have attended; like weekends I'm not usually very busy. MT100 tel |
|  | You should have clinics on Saturdays because that is when most people are not held up at work. Having the clinics on Monday is hectic as people are held up at work and getting permission is quite difficult... Sometimes bosses do not grant day offs on busy days, especially week days. If you could make the clinics to be on a weekend many people will attend, I will also attend if you make the clinic day to be on a weekend. When you make it on a Saturday many people will attend because even if they are busy they get off from work quite early at around 1pm. MT17 – tel<br>I am always free on Saturday and Sunday... I think that Monday, Wednesday, or even the whole week of clinic will be easier for most people to access the clinic as it is easier to ask for permission from work and be sure you will be attended. With the whole week one can get a day to go and if they do not get attended that day they can still chose the next day and will be sure to be attended. MK040 – tel<br>Maybe if the exercise could be scheduled off working days - that's during the weekend, in Saturday or Sunday, because at that time am free without work I could turn up. MFK02 – in-person |
| Add more clinic locations | Have more many work station where people can easily access the services to reduce the distance people need to travel to get treatment. Even the elder people can reach the outreaches also it give them option on where they can get treatment because on line on a particular station but be short where as the other might be long. MT 010Z – tel<br>Some people should be referred to other hospitals like MTRH. MT48 – tel<br>Have eye medics even in Nyeri. MT984 – tel<br>Next time there such a project, its best if you people divided the community people according to their villages or maybe |
|  | sub-location 9:10 so that people don't overcrowd in the same place like that day because I think that day the whole location was referred to Kambiti which made the area overcrowded and we could not receive the services because the people were many. MT38 – tel<br>Set more clinic centers so that people will not need to travel from far. MFK2 06 – in-person<br>You should have many stations and also the doctors attending people should be many. Some other places can be used like churches and it will be easier for people to go to access the place. MFK009 – in-person<br>You know so many people have eye problems like all those that were that day. Next time have many centers, like three, not one like that day. MFK04 – in-person<br>Let there be more facilities for the clinics, like five stations. That would be OK. MFG07 – in-person<br>If there will be more stations for check-up that would ease the long queues. Even the clinics should be set up even in schools nearby, then people would not be as many that day. At least five stations would be good in this area of Gakoromone. Come with machines even to the nearby schools and set more stations, and also doctors to be added in those stations. MFG05 – in-person<br>For the long queues, am suggesting if there will be more screening centers to avoid overcrowding at one center and the long queues. MFG03 – in-person |
| Mop- up clinics | The outreach should be conducted another time so that the people who were left unattended could be attended. MT48 - |
|  | tel<br>If the outreach is done once more I could go. You should come back because there more people who were not attended to - MT47 – tel<br>Just tell us when you are coming back to the community for other clinics. Even this week you can tell us you are coming and we shall attend... Plan for when you are coming next, three days this time will be better so that if someone misses one they can still have a chance. MT496 – tel<br>Let's say, those people who never managed to attend on that particular appointment day, they should be followed up immediately maybe later that day or the following day so that their appointment can be rescheduled. For me, I would have wished that the exercise would be repeated for more days following up on those who missed the opportunity. MFK01 – in-person |
| Give each person a specific appointment slot | The attendance number in the outreach should be issued in line with the referral messages clients received. The message should also have the attendance number and the time allocation to help the clients organize themselves to attend. MT772 - tel |
| Schedule fewer people to attend each day | I think you should schedule less people. MFK04 – in-person<br>I would also like it if you control the number of people you schedule for a particular day, because now you have experience of how many people you can attend for a day, schedule just that amount of people so that everyone may be attended to. That way everyone will be aware what time they will attend so that they may not waste the whole day. It wasn't fair of you to call that many people, they were already sick and they had to wait for hours under the scorching |
|  | sun. MFK 01 – in-person |
| **Awareness & Communications** |  |
| SMS reminders, including on the day of the clinic | Sending them SMS message and reminding them to attend their clinic appointment or calling them. MFK008 – in-person<br>That same same day for the clinic I should receive a reminder message to remind me to attend. I have many things I can easily forget... A reminder message should be sent before and during that day to attend the clinic. MT656 – tel<br>They should make announcements early; not a day before the clinic.. They can make announcements three days before so that you can be prepared. Even sending messages four days before, its OK. MFK2 05 – in-person<br>Next you send me a message to remind of the clinic and a phone call will be okay to remind me of the appointment. MT36 - tel |
| Send a text to individuals who missed the outreach clinic to invite them to attend on another day | If a person never made it to the outreach, he should be notified of the next eye clinic outreach even if it is in another location. Because one cannot be busy all the time. MT46 - tel |
| Phone call reminder, especially for those who cannot read | Next you send me a message to remind of the clinic and a phone call will be okay to remind me of the appointment. MT36 – tel<br>Sending them SMS message and reminding them to attend their clinic appointment or calling them. MFK008 – in-person<br>If someone didn't make to attend the outreach, you can call the to remind them. MFNG03 – in-person |
|  | Maybe you can remind us by calling us as you have today. MFKI08 – in-person<br>In order to create the trust and be sure that this exercise is medical, at least you should call on the day of the check-up so that you can explain the exercise in detail - so that we can get to trust the exercise because sending just a message as a reminder is not convincing enough or satisfactory. MFI 04 – in-person |
| Explain why attending clinic is important at the point of referral | Also during screening we should be given more information why this clinic is important. MT656 – tel<br>They should also be informed the importance because some people may end up blind. MFNG03 – in-person |
| Use churches and radio broadcasts to remind people to attend | Mobilize the clinical outreach especially on churches and radios. MT48 – tel<br>Also, the activity should be announced and advertised to people in public, for example in TV or radio shows. At least even if I don't know you, someone else might have seen you, so people will be able to come more freely without the fear of being scammed or conned. MFI 04 – in-person<br>It should be announced in public so that everybody will get there because the message can come when the phone is off. Let it be publicly announced that certain day will be clinic day, we meet at a specific place. MFG 07 – in-person |
| Specify clinic dates and times | I would propose that if you have a project like that one you communicate well on the exact opening and closing times, so that people may not come and get stranded outside and eventually go home without getting treated. There were so many people outside the facility who got devastated because they thought that the exercise would go on at least up to |
|  | close of business hours. I feel like if you communicated well on the closing hours I would have programmed myself well and come in the morning whereby I could get the treatment that I required. MFNG03 – in-person<br>You should communicate on the time because I did not know that the clinic was opening early, and tell the people to come early enough so that they may be attended effectively. MFK04 – in-person<br>Communicate well on how many days you will be available, when I know the duration which you are staying I will avail myself. MFK03 – in-person |
| Allow people to choose their appointment day | Next time they should ask me when I am available instead of putting a date for me because of my work. The biggest challenge for me was timing. MT100 – tel<br>Communicate with client if they will be available for the outreach. If not allocate them another day and time. MT894 – tel |
| Clarify what services are available at the outreach clinic and specify if there are any costs linked to these services | Clarify information because of the Worldcoin thing, so during screening we should be given more information. Anything free people think its hidden things. MFK2 06 – in-person<br>People should also be announced that they will be helped, not only will they be screened, there will be other help. MFNG03 – in-person<br>They should tell me that the services are free so that ill not assume that there are charges... they should tell us the clinics are free. MFK2 04 – in-person |
| **Norms, values & health beliefs** |  |
| Sperate younger people, women & children, and elderly people | Address the line issue and have women on their [own] lines. But youth can just have their line where we can mingle and interact - for example on how hard the economy is - and by the time we realise the line is shortening. But with women on the same line we will not have anything to talk about. MT040 – tel<br>One day of the outreach should be reserved for the elderly and the other day for the other people since you should give priority to elderly people. If you allow everyone to the outreach there will be long queues and many of them will not be attended. MFKIA 012 – in-person<br>What I would want you to improve is that next time hire competent queue managers who control how people are coming in, and the elderly, pregnant mothers and mothers with small children to be put on their own queue so that there will be effective treatment. MFK01 – in-person |

## Appendix 7: Original drafts: Intervention counselling script and SMS reminder

### Original enhanced counselling script

I have found a problem with your eyes. I am referring you to the outreach treatment clinic that will be held at [location] on [date] between [time] and [time]. At the clinic, eye care professionals will perform a specialist assessment and provide medicines and spectacles as required, all free of charge. Note that a small proportion of people who attend the clinic will be found to have complex eye problems that require onward referral for hospital assessment and specialist glasses. This may incur a cost. However, the vast majority of people have their needs fully met for free at the outreach triage clinic and do not require any further referral.

With treatment, you will be able to see more clearly. This will help with your work, reading, viewing screens, and many other things. It is important that you attend the clinic or your eye problem may get worse. The clinic will only be running from [day] to [day], so if you don’t manage to attend, you may not be able to get free care again in the future.”

### Original intervention SMS script

We found that you had an eye problem. Please attend the outreach clinic at <> on <> between 9am-5pm to receive free medicines/spectacles If you are found to have a complex eye problem, you may be referred to a hospital for further care or specialist glasses, and this may include a fee However, the vast majority of people who attend the outreach get their eye problem fixed, for free, without the need for any further referral It’s important that you attend, as your eye problem may get worse, and you might not have a future opportunity to access free care. See you on <

### Changes

We made three changes based on feedback:

1. One of the young men from the left behind group wanted us to change the sentence *“It is important that you attend the clinic or your eye problem may get worse”*, to make it ‘less scary’. We had included this original sentence based on interview feedback that we needed to better explain the importance of attending. For instance: “They should also be informed the importance because some people may end up blind”. We changed the wording to read; “It’s important that you attend to protect your vision.”
2. Due to inflation-imposed budget constraints, the clinics are no longer able to provide all glasses and medications for free. At this point in time it is not clear how subsidised the costs will be. We removed wording stating that the treatments will be free. We kept wording stating that the assessment will be free as this still represents a major advantage over attending a high-street provider or hospital clinic.
3. Representatives from Peek vision advised that “see you on <>” might be misleading, as the day of the appointment might be more than a week away (e.g. next Friday rather than this Friday). We changed this auto-populated section to “see you on <>”.

## Appendix 8

Barrier and solution quotes from the interviews with high-income people.

Appendix 8: Barrier and solution quotes from the interviews with high-income people
| Themes | Quotes |
| --- | --- |
| **BARRIERS** |  |
| **Costs** |  |
| Perceived cost of drops/spectacles/services | - There is a friend of mine who was also referred to general hospital to buy specs which were costing 3500 shillings [7.37]. (HIG7)<br>- I just know if I come for the clinic, I will be told I have a problem and I will be required to pay for the services but if I know I will get free services I will be motivated. (HIG4) |
| Loss of wage/income | - N/A |
| Transport cost. | - The day of appointment I got a job transfer from Meru to Maua (2:50). It's the transport from Maua to Meru the fare is too high (HIG3) |
| **Distance and transport** |  |
| Distance to the referral site | - N/A |
| **Desire and priority to seek care** |  |
| Conflicting work engagement | - I was given a date to be checked but that day I went to work in Nairobi, we had other programmes in Nairobi that day on insurance that's why I did not make it [2.35], I work with insurance and I often go to Nairobi for work, I don't have specific time or date to go to Nairobi but mostly I usually go on Thursdays and Mondays [4.30]. (HIG1)<br>- I didn't attend the clinic since I travelled to Isiolo and that day I was working in Isiolo, [3.10]. |
|  | It was a working day, and I was on duty, I didn't get some time to attend the outreach clinic. (HIG2)<br>- The day I was supposed to go for clinic i went back at work in Nyeri. The day of clinic appointment was the same day i went back for work. (02:42) (HIG5)<br>- There is a place I went that day, I went to work in Naivasha, I don't work specifically in Meru, the work I do is usually anywhere like now I am in Maua [1.05], I do fitting, I fix things like gypsum and wardrobes. I can come another day especially from next week because now I am free [2.25]. (HIG6) |
| Other conflicting engagement | - I was screened and told that my left eye had a problem, was referred to Kinoru dispensary but I was sick that day. That whole period I was unwell and that specific day I was weaker (HIG4)<br>- I was told to first buy a certain drug and then go to general hospital in case I don't improve [1.15], after using that drug the problem cleared so I dint go to the general hospital. I had told them I wouldn't get time to go to the hospital they told me to buy that drug I use before I organise myself [4.00]. I used that drug for just one month but the problem cleared totally (HIG7) |
| Forgot | - I was screened and told that my left eye had a problem, was referred to Kinoru dispensary but I was sick that day and I also forgot about the clinic [1.43]. (HIG4) |
| **Clinical service quality** |  |
| Long queue | - I went and found that the queues were very long at Kinoru dispensary (2:23) I went around 10am.i stayed for about an hour. And I work at Makutano, so I went back. (HIG3)<br>- The queue was too long, people were so many, so I just went back to job [0.52], where they |
|  | were offering the service was different from where we see patients, we were not sharing the clinics we had given them a separate place but within the same compound [1.35]. (HIG8)<br>- Reason why I did not go is that I used my card and I decided to go another hospital and use my card instead of Meru general hospital, where I was referred. I went to life care hospital. Even if I go to general ill find long queues (11:30) (HIG10)<br>- I went around 3 pm and the queues were very long around 2000 people many people did not go to see doctor due to long queues still at 3pm there were a lot of people (4:34) then i decided to go back home (HIG11)<br>- I was worried about going to the hospital because there is usually so much queue (HIG7) |
| Insufficient numbers of staff | - N/A |
| **Facilities** |  |
| Clinic not open at stated times | N/A |
| **Awareness & communication** |  |
| Lack of clear information on services available | - N/A |
| Lack of clear information on appointment days | - We were not told the specific day to go to the hospital, maybe we can be told to go a specific day and be given a note so that one can know that a certain day he/she will be going to the hospital. (HIG7) |
| Lack of clear information on appointment times | - N/A |
| Insufficient counselling | - N/A |
| Did not receive reminder message | - N/A |
| **Fear** |  |
| Fear | - N/A |
| **Norms, values, health beliefs** |  |
| Discomfort with mixed genders in the queue | - N/A |
| Male health seeking behaviour | - N/A |
| Assumption that eye problem wouldn't be addressed | - N/A |
| Dislike crowded places | - N/A |
| Sought services elsewhere | - N/A |
| **SOLUTIONS** |  |
| **Costs** |  |
| Provide transport fare | - Provide us with transport, if provided with fare I can come, 730 shillings will be okay for me [15.12], I can request for permission to go to the clinic for few minutes because people get permission even to go to hospital when they fall sick, [15.39] (HIG2) |
| Subsidise treatment | - If you could have convinced me and promise me that I would get spectacles for free, I could get motivated [8.15]. I just know if I come for the clinic, I will be told I have a problem and I will be required to pay for the services but if I know I will get free services I will be motivated. (HIG4) |
|  | - I wouldn't even forget to attend the clinic [8.24], for example if you offer things like operation for free or even lower the cost of the services then people would be so motivated (HIG4)<br>- That drug was so costly, I bought it at 1050 at [location] chemist [4.17], many people would not afford to buy drugs so in future you can have variety of drugs and even different spectacles even if we can pay for some amount, you get served once [7.50]. There is a friend of mine who was also referred to general hospital to buy specs which were costing 3500 shillings [7.37]. (HIG7) |
| Pay people to attend | - N/A |
| **Distance and transport** |  |
| Move clinics closer to where people live and work | - Since you people are concerned and are following up on us, you can even come to the remote areas for those who can't access the clinics in town since there are so many people with eye problems, like the way CHVs works, you can even take people like me to go round to remote areas since there are mothers who can't reach the clinics in towns. you can also get to other places like Samburu rather than proving services only to Meru County, it was even in media that a place like Samburu people have eye problems so you can visit such places as you observe what people are going through, [9.12] (HIG2)<br>- If I can do my appointment where I am e.g. Nyeri or Kirinyaga, if the service could be brought near my working area (04:44) (HIG5)<br>- Bring services closer to people especially in rural places (4:48) in a school or a public ground (HIG9) |
| Provide transport | - N/A |
| Avoid setting up clinics in areas with difficult terrain | - You should also avoid rural areas where roads are not accessible e.g. that time it was raining and therefore most of the people wouldn't make it to the clinic, [7.40]. (HIG1) |
| Door-to-door services | - N/A |
| **Clinical service quality** |  |
| Improve punctuality and speed of staff | - N/A |
| Ensure that all medicines and glasses can be provided on-site | - N/A |
| Provide high quality spectacles/frames | - Next time come with spectacles of good standard, not for the old people. Come with glasses with good standard (6:39) (HIG3)<br>- ...and next time come with spectacles of different types and medication (HIG10) |
| Add more staff at each clinic | - Adding the number of medical staffs so that people can be attended fast, if I had gone to the clinic and found a long queue I would just go back since I can't keep waiting in the queue, If someone come from far and the services are given fast, he/she can't feel like they are wasting time. (HIG1) |
| Schedule fewer people to attend each day | - if they do it again, they say a particular area should be attended at a particular day this will avoid overcrowding and long queues... (HIG10) |
| **Facilities** |  |
| Keep the clinics open for a greater number of days | - Adding the number of days to be twice a week or if its monthly once a week (HIG8) |
| Extend clinic opening hours | - N/A |
| Keep the clinics open on the weekend | - If it was on weekend Saturday I would attend (03:21) (HIG5)<br>- you should do the clinics during weekends especially Saturdays [4.34], a day like Wednesday is very tricky, most people work the whole week then get paid on Saturdays so they can make it to attend the clinic [5.24]. (HIG6) |
| Additional workstations | - many stations to see people and to have many doctors (5;15) (HIG11) |
| Add more clinic locations | - You can set up other places for the outreach clinic like Nairobi and other places for those travelling during that day [5.40] give many places where people can be checked for example the way I didn't make to attend the clinic in Meru I could have found some few hours to be checked somewhere else, [5.40]. (HIG1) |
| Mop- up clinics | - Schedule another clinic and I will come, inform me like one week before so that I will be |
|  | prepared [3.01]. (HIG6) |
| Give each person a specific appointment slot | - Give people different appointment dates to avoid overcrowding (7:57) a particular number of appointment for a specific day of appointment (8:08) and come with a message to show u are supposed to be here today (8:17)dont give all people one day for appointment. (HIG3) |
| Schedule fewer people to attend each day | - N/A |
| **Awareness & Communications** |  |
| SMS reminders, including on the day of the clinic | - You should also send text messages like the way you sent me a message last time [10.03] (HIG2)<br>- Remind people about the clinic before the clinic day by sending text messages...Even though I was sick, if I could be reminded, I would try my best to come. (HIG5)<br>- Send people text messages twice in the phone like one day before the clinic [10.38]. (HIG6) |
| Send a text to individuals who missed the outreach clinic to invite them to attend on another day | - N/A |
| Phone call reminder, especially for those who cannot read | - N/A |
| Use CHPs to remind people to attend | - The CHVs can be going home to home telling people 'This is the day you should come to see the doctor' [4.26]. (HIG8) |
| Explain why attending clinic is important at the point of referral | - ...also educate people about its importance as you convince them, also extra services like spectacles [6.09]. (HIG5) |
| Use media, posters, churches and radio broadcasts to remind people to attend | - Advertise using posters and media so that people can get the information, during the screening not everyone was aware and not all were screened, other people were not around so if you advertise everyone gets the information, [10.02]. (HIG1) |
|  | - Advertise more like using stickers where there is movement of people like Makutano [9.35] some people didn't know about the screening and others also forgot [10.04] (HIG6) |
| Specify clinic dates and times | - N/A |
| Allow people to choose their appointment day | - N/A |
| Clarify what services are available at the outreach clinic and specify if there are any costs linked to these services | - N/A |
| **Norms, values & health beliefs** |  |
| Separate younger people, women & children, and elderly people | - N/A |
| Other | - I can request for permission to go to the clinic for few minutes because people get permission even to go to hospital when they fall sick, [15.39]. (HIG2) |

